# Harmony COVID-19: a ready-to-use kit, low-cost detector, and smartphone app for point-of-care SARS-CoV-2 RNA detection

**DOI:** 10.1101/2021.08.12.21261875

**Authors:** Nuttada Panpradist, Robert G. Atkinson, Michael Roller, Enos Kline, Qin Wang, Ian T. Hull, Jack H. Kotnik, Amy K. Oreskovic, Crissa Bennett, Daniel Leon, Victoria Lyon, Peter D. Han, Lea M. Starita, Matthew J. Thompson, Barry R. Lutz

## Abstract

RNA amplification tests allow sensitive detection of SARS-CoV-2 infection, but their complexity and cost are prohibitive for expanding COVID-19 testing. We developed “Harmony COVID-19”, a point-of-care test using inexpensive consumables, ready-to-use reagents, and a simple device that processes up to 4 samples simultaneously. Our lyophilized reverse-transcription, loop-mediated isothermal amplification (RT-LAMP) can detect as little as 15 SARS-CoV-2 RNA copies per reaction, and it can report as early as 17 min for samples with high viral load (2 x 10^5^ RNA copies per reaction). Analysis of RNA extracted from clinical nasal specimens (n = 101) showed 95% concordance with RT-PCR, including 100% specificity in specimens positive for other viruses and bacteria. Analysis of contrived samples in the nasal matrix showed detection of 92% or 100% in samples with ≥20 or ≥100 particles per reaction, respectively. Usability testing showed 95% accuracy by healthcare workers operating the test for the first time.

**ONE SENTENCE SUMMARY:** Harmony COVID-19: point-of-care SARS-CoV-2 RNA detection

## INTRODUCTION

In 2019, an outbreak in China of SARS-CoV-2, the causative pathogen of COVID-19, rapidly became a global pandemic(*1*), and after a year it has infected 100 million people and killed 2 million people(*2*). Multiple measures have been used to contain the spread of COVID-19. Governments imposed universal "stay-home" orders to minimize transmission(*3*) which in turn has harmed mental health, social life, and the economy(*4*). It has been proposed(*5*) and demonstrated(*6*) that widespread COVID-19 testing and contact tracing could be a solution for re-openings, allowing subsets of a community to resume work and begin to restore the economy. Moreover, testing is essential for re-opening of international borders. Several countries have enforced a “fit-to-fly” policy; international travelers must test negative for COVID-19 within 72h before boarding international flights(*7*). Fast and sensitive point-of-care (**POC**) testing could facilitate the safe return to functioning domestic and international economies. POC testing could allow testing to expand to geographic regions that have limited access to centralized laboratory facilities, enable essential businesses to regularly test employees, and allow easy and efficient community testing by public health officials and clinics. Importantly, POC testing is critical in settings that require rapid turnaround time such as emergency urgent care facilities or points of introduction such as airports.

Here, we report the development of Harmony COVID-19 — a complete moderate-throughput sample-to-result system for sensitive POC detection of SARS-CoV-2 RNA. Harmony simplifies testing with ready-to-use assay reagents, an easy-to-use dedicated smartphone interface, and an inexpensive isothermal heater/detector device to enable POC testing. The assay includes three redundant SARS-CoV-2 targets to avoid false negatives due to viral mutation and an internal amplification control (**IAC**) in each test to avoid false negatives in case of assay failure. The heater/detector detects two-color real-time fluorescence for the SARS-CoV-2 redundant targets and IAC, and results are automatically reported on the smartphone. We evaluated the system using two panels of specimens - a panel of extracted nasal specimens from individuals with respiratory symptoms, and a blinded panel of contrived specimens from the XPRIZE Rapid Covid Testing competition – and we conducted usability testing of Harmony by healthcare workers (**HCWs**). These evaluations have demonstrated that Harmony is a complete system for sample-to-result testing that is high performance and sufficiently simple for CLIA-waived settings.

## RESULTS

### Workflow and operation of COVID-19 Harmony

The Harmony test kit and detector were engineered for low cost and simplicity of use to enable POC testing (**Fig.1a**). The test involves taking a nasal swab, eluting the swab in a lysis buffer, and transferring the buffer to a reaction tube containing ready-to-use reagents. The reagent tube is inserted into the custom-designed heater/reader operated by a cell phone that provides instructions and displays the result.

The Harmony assay uses a unique variation of reverse transcription loop-mediated isothermal amplification (**RT-LAMP**) that allows fluorescence detection of multiple targets in the same reaction. The assay amplifies three genomic regions of the SARS-CoV-2 nucleocapsid phosphoprotein reported by a green fluorescence signal (FAM) and an engineered internal amplification control (**IAC**) reported by a red fluorescence signal (TEX 615, alternative for Texas Red®). For samples with no target (**Fig. 1B**, left; NTC), detection of the IAC confirms that the reaction was functional; absence of target and IAC indicates a failed reaction. For samples with SARS-CoV-2 RNA (**Fig. 1B**, middle; 200 copies/rxn), the green signal indicates a positive test, regardless of whether IAC is detected. Harmony using dry reagents detects down to 200 copies of SARS-CoV-2 RNA on a swab (**Fig. 1B**, right; ∼20 copies/rxn, corresponding to 0.5 copies/*µ*L). Real-time detection reports a positive sample as soon as the target signal appears, allowing detection in <30 min for samples ≥2000 copies per swab (**Fig. 1C**), with earlier results for higher viral load. The three SARS-CoV-2 targets are reported into the green fluorescence channel to provide a triply-redundant assay to make it robust against viral mutations (**Fig. 1D**). A positive test result is reported when any of the three SARS-CoV-2 targets are present, and a negative test result is reported if SARS-CoV-2 RNA is undetected, and the IAC is detected. If neither target nor IAC is detected, it indicates a test failure, and the result is reported as indeterminate (**IND**). **table S1** lists the sequences of primers/probes used in this RT-LAMP assay.

**Fig. 1.**
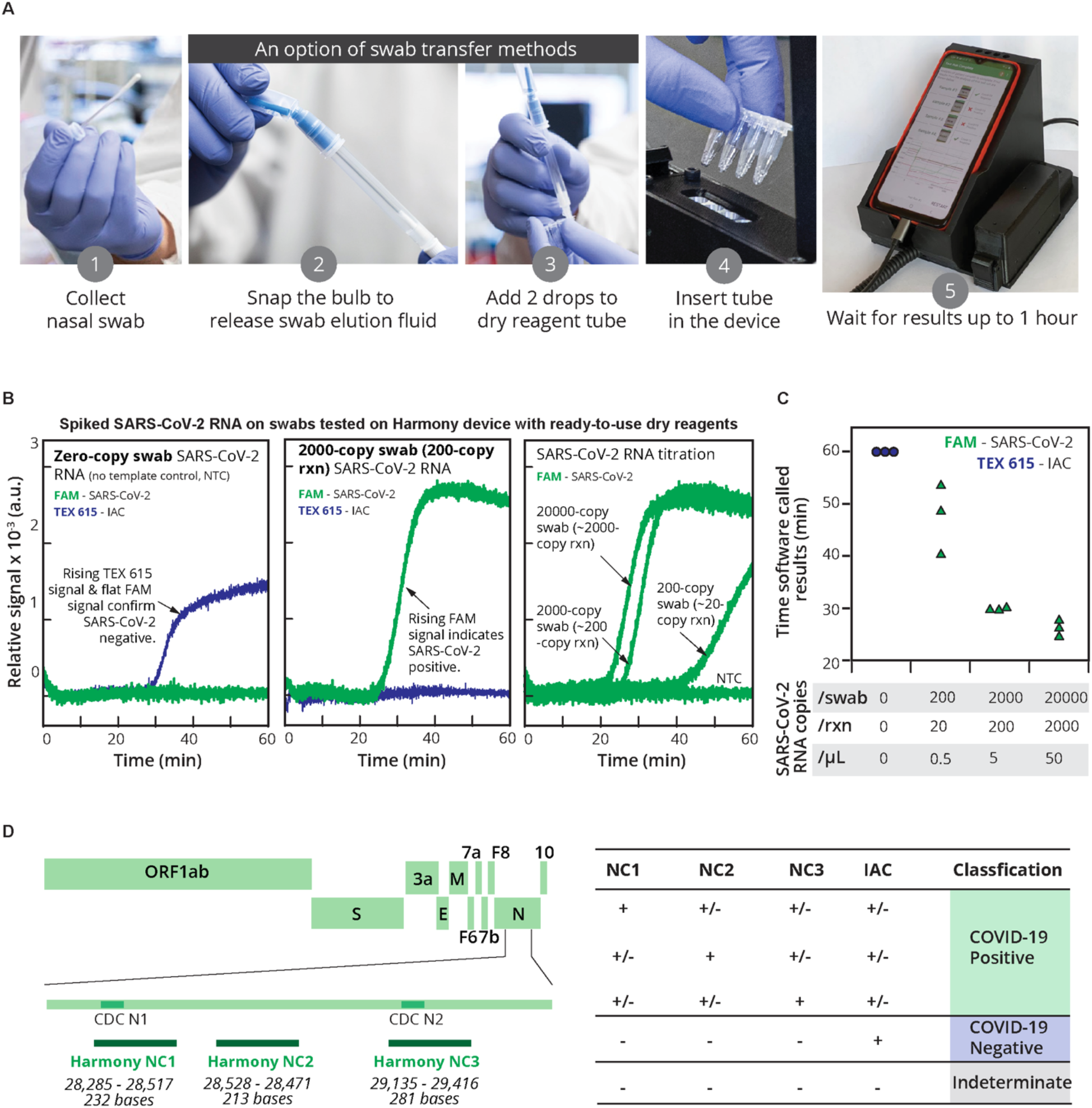
Harmony COVID-19 workflow, analytical performance, and interpretation. (**A**) User workflow. A nasal swab is collected and eluted in the rehydration buffer using either a unified sampler-dispenser (shown) or a tube-and-bulb method (**Fig. 6**). The swab eluate is then transferred into the reaction tube to rehydrate lyophilized, pre-loaded RT-LAMP reagents. Users follow the instructions on the cell phone app to record the sample identity, insert the tube in the test slot, and close the device. Four samples can be analyzed in parallel. (Photo credits: a1-a4, Mark Stone at the University of Washington and a5, Christopher Snyder at North Seattle College). (**B**) Examples of on-device analysis. Lyophilized reagents were rehydrated with the eluate from swabs spiked with 0, 200, 2000, or 20,000 copies of SARS-CoV-2 RNA, run on the device, and analyzed in real time. The samples with SARS-CoV-2 RNA at 2000, 200, and 20 copies/reaction (**rxn**) were reported as positive by software at 27 min, 30 min, and 41 min, respectively; after 60 min the NTC reaction was classified as negative. (**C**) Time-to-result. Individual detection time points (*n = 3*) are plotted. Variation in detection time increases as samples approach the detection limit. (**D**) Diagnostic algorithm. Harmony COVID-19 software calls samples as positive, negative, or indeterminate based on detection of the three regions in nucleocapsid gene (NC1 overlapping with the CDC N1 region, NC2, and NC3 overlapping with the CDC N2 region) and the engineered IAC sequence.

### Development and analytical sensitivity of ready-to-use reagents

We have developed a lyophilized RT-LAMP mixture that contains primers, sequence-specific fluorescence probes, IAC DNA template, polymerases, supporting proteins, and co-factors (**Fig. 2A**) and is activated by adding the nasal swab eluate containing additional buffer, salts, and detergent. We initially screened excipient formulations that can preserve enzymatic activity and do not significantly interfere with the primer or probe behaviors. Mannitol did not drastically impact the core assay compared to trehalose at the same concentration (**fig. S1**) and did not require increasing the reaction temperature to maintain the assay speed. Using a commercial real-time thermal cycler, lyophilized RT-LAMP detected down to 15 RNA copies/rxn (0.38 copies/*µ*L), and IAC was detected in all negative samples (**Fig. 2B**). RT-LAMP amplification of 2x10^5^ RNA copies/rxn using the dry reagents generated detectable signal in 17 min (**Fig. 2C**). We subsequently tested our in-house lyophilized reagents on a larger number of technical replicates of SARS-CoV-2 RNA at 20 copies/rxn using multiple batches, and 90% (36/40, 95% confidence interval (**CI**): 76-97%) were amplified (**Fig. 2D**). Medians of time to detection for FAM and TEX 615 signals were 28.6 min (range: 28.0-29.4) and 31.7 min (range: 31.1-32.7), respectively. In the presence of simulated nasal matrix (salt, mucin, human DNA), we observed a 10-min delay in amplification of both SARS-CoV-2 RNA and IAC DNA (**Fig. 2E**), but all targets were detected correctly. Greater interference or other harm to the reaction will further delay IAC amplification; thus, the IAC serves as an indicator of the reaction’s function and can prevent reporting false negatives due to interference or damaged reagents. By setting a 1-hour reaction time, the assay can suffer some delay while still correctly identifying positive samples. Specificity testing against closely-related pathogens MERS and SARS-CoV-1 showed no cross-reactivity (fig. S2).

**Fig. 2.**
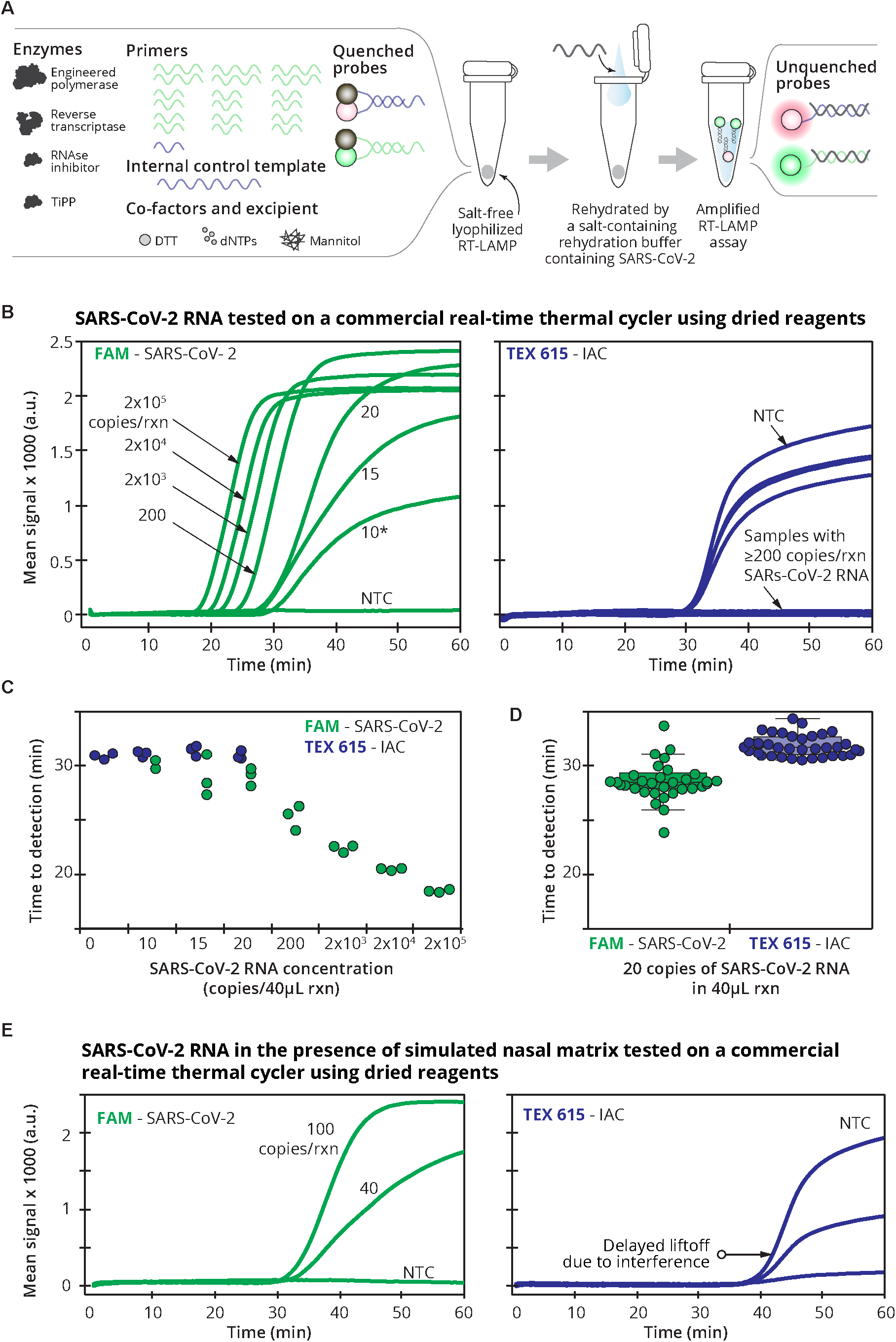
Lyophilized RT-LAMP and its analytical sensitivity. (**A**) To set up the assay, a sample in elution/rehydration buffer containing magnesium, Thermopol buffer, and detergents is added to rehydrate the lyophilized RT-LAMP reagents in a single step. (**B**) Real-time amplification signal of RT-LAMP reactions containing SARS-CoV-2 RNA (**NTC**; no template control), 10, 15, 20, 200, 2000, 2x10^4^, or 2x10^5^ copies/40µL reaction (**rxn**). The left panel shows the average (*n =3*) FAM signal detecting SARS-CoV-2. *Only 2/3 replicates of 10 copies/rxn amplified so the average was from duplicate reactions. The right panel shows TEX 615 signal detecting the internal amplification control (IAC). The IAC was detected in the NTC (*n* = 3) and in the single 10 copies/rxn that did not detect SARS-CoV-2. (**C**) Time to detection of SARS-CoV-2 and IAC targets reported by the real-time thermal cycler. Individual data points are plotted. Note that only 2/3 replicates of 10 copies/rxn amplified. IAC signals were properly detected in NTC (*n = 3*) and 1 of the 3 replicates of 10 copies/rxn undetected for SARS-CoV-2. (**D**) Evaluation of analytical sensitivity at 20 copies/rxn of SARS-CoV-2 RNA (*n = 40*), performed in two different runs (*n = 20* each) with two different serially diluted RNA samples. Time-to-result of individual samples are plotted along with medians (middle lines of the boxes), interquartile ranges (edges of the boxes), ranges (the ends of whiskers), and outliers (data points outside the whiskers). (**E**) Impact of nasal matrix. SARS-CoV-2 RNA at 0, 40, or 100 copies/rxn was amplified in lyophilized RT-LAMP in the presence of simulated nasal matrix (*n = 3*). All data was measured every 13s using FAM and TEX 615 channels, by a real-time thermal cycler in 1h reactions at 63.3°C, and (**C**).

### Hardware: housing and real-time fluorescence reader device

The Harmony system includes a heater/detector device controlled by a dedicated cell phone and plastic housing with a sample setup station (**Fig. 3A**). The heater/detector device (**Fig. 3B**) includes an aluminum heater block with four wells and a spring-loaded heated lid to prevent condensation. A proportional integral derivative (**PID**) feedback loop running on a microcontroller allows recovery of the temperature setpoint within 3-4 min after disturbance from opening the lid (**Fig. 3C**). Two LEDs (**Fig. 3D**) provide the excitation light by shining down through the top of each sample tube. Fluorescence emission passes through holes drilled on the side of the heater block. Each sample well has an emission filter/photodiode set on each side to detect two emission wavelengths (**Fig. 3E**). A blue LED (dominant wavelength 470nm) is used to excite FAM, and the emission photodiode sits behind a dielectric 550nm longpass filter. A yellow LED (dominant wavelength 587nm) is used to excite TEX 615, and a second emission photodiode sits behind a 630nm longpass dielectric filter.

**Fig.3.**
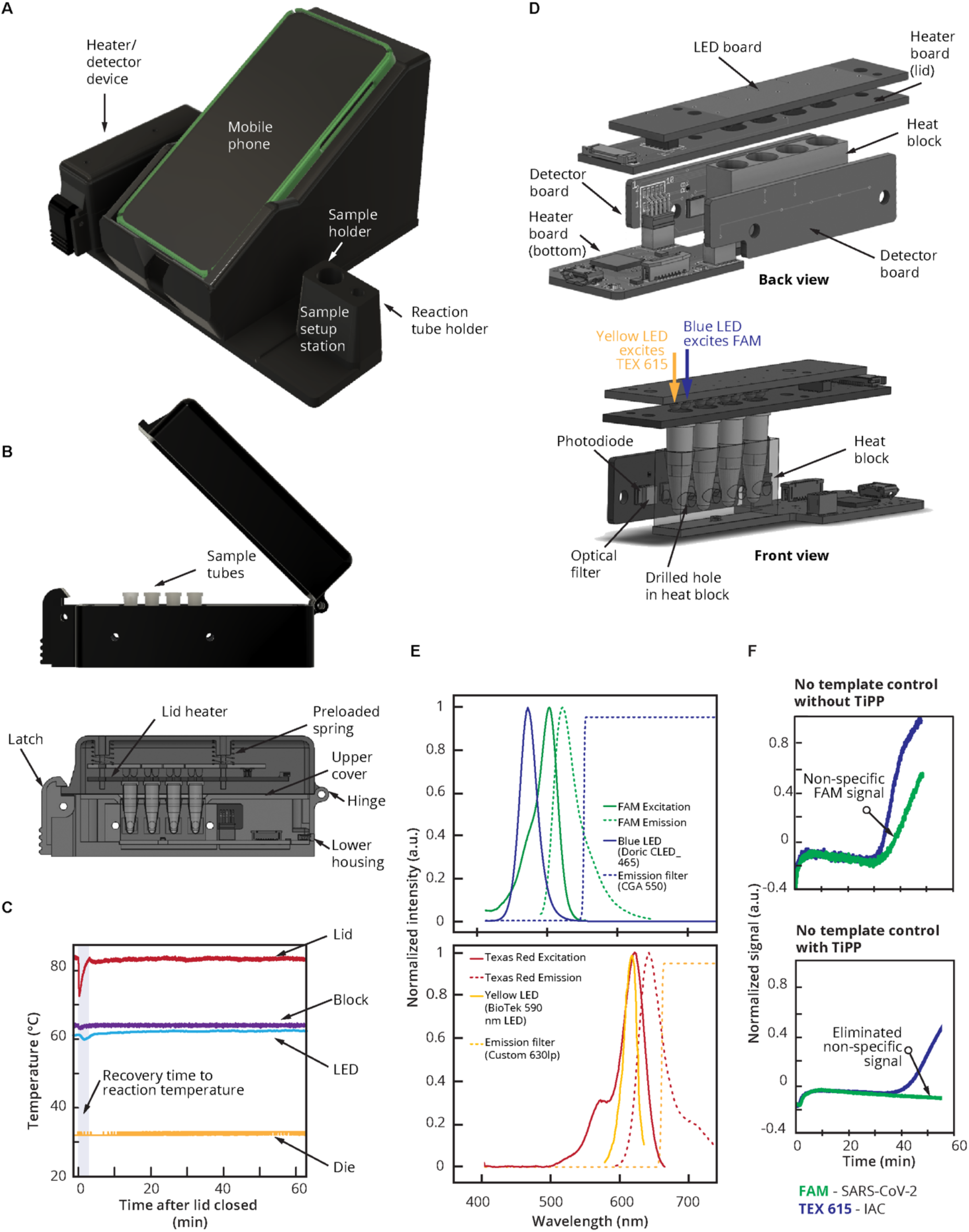
Overview of Harmony COVID-19 hardware. (**A**) Plastic case cradles the cell phone (removable and connected by a spring cable), mounts the heater/reader device, and houses a Charge-Plus USB-C connector hub (not shown). A sample station holds the sample and reaction tube during processing and is detachable to allow cleaning. Weight: 266g housing, 104g heater/reader device (total <0.5kg). Dimensions: 205mm x 167.5mm x 125mm housing, 30mm x 120mm x 50mm heater/reader device. (**B**) The heater/reader device contains an aluminum block with wells for four samples (0.2mL reagent tubes) and is heated from the bottom by a resistive heater made from serpentine conductive traces on a printed circuit board. In the middle of the heater, a temperature sensor (Class B Platinum RTD) is used to measure the heat block temperature. Power to the heater is supplied by a MOSFET and modulated by varying the PWM duty cycle of the MOSFET. (**C**) Temperature profile after the lid opening throughout a 60-min assay run. The microcontroller (die) is measured to ensure the electronics are not overheated. (**D**) Lid heater, LED, and detector. A heater (85°C) on top of the tube prevents condensation on the tube lid. The lid heater is spring loaded to apply a preload force to the tubes to ensure good thermal contact and contains pass-through holes for excitation LEDs. (**E**) Spectral characteristics of the detection system. Blue and yellow LEDs provide excitation for FAM and TEX 615, respectively, and photodetectors on each side detect light passing through emission filters (spectral information from manufacturers(*21–23*)). The LEDs are operated independently (only one LED is illuminated at a given time). (**F**) Effect of thermostable inorganic pyrophosphatase (**TiPP**) on RT-LAMP signal in the Harmony device. Normalized signal of negative control samples using the RT-LAMP formulation with and without TiPP (*n = 1* each) at 64°C.

### Thermostable Inorganic Pyrophosphatase enables real-time LAMP detection in the Harmony COVID-19 device

RT-LAMP generates inorganic pyrophosphates that precipitate from the solution. The cloudiness created by precipitates has been used as a visual readout signal for LAMP(*8*), but here the cloudiness scattered light and caused non-specific signals in NTC samples (**Fig. 3F**, top) when tested on device. Adding inorganic pyrophosphatase in the RT-LAMP reaction eliminated built-up pyrophosphate and prevented non-specific signal in negative reactions (**Fig. 3F**, bottom). Interestingly, this effect was not observed when the reaction was tested in the commercial thermal cycler (BioRad CFX96).

### Multi-purpose Android mobile app: instruction, temperature control, and real-time analysis of results

The phone software provides interactive guidance for setting up the test. It prompts the user to input a sample ID by scanning a barcode, capturing a photo, or typing a sample ID (**Fig. 4, A-D**). The software also carries out the closed-loop temperature control for the heater block and lid heater, and it displays the status of the heater block to prompt the user to insert the reaction tubes only after the heater reaches the reaction temperature (**Fig. 4E)**. During the test run, the screen displays the test status (“analysis in progress”) and elapsed time, or it reports any detected errors (“failed run”) (**Fig. 4F**). Importantly, the software performs real-time data analysis that detects fluorescence intensity rise above a dynamically and automatically computed background level. Signals from SARS-CoV-2 can appear early in the test, especially for high viral loads, and the real-time analysis allows reporting of positive tests immediately after they are detected (**Fig. 4G**). Negative results and IND results are reported at the end of the test run time (**Fig. 4H**). Software analysis is described in the Methods section.

**Fig. 4.**
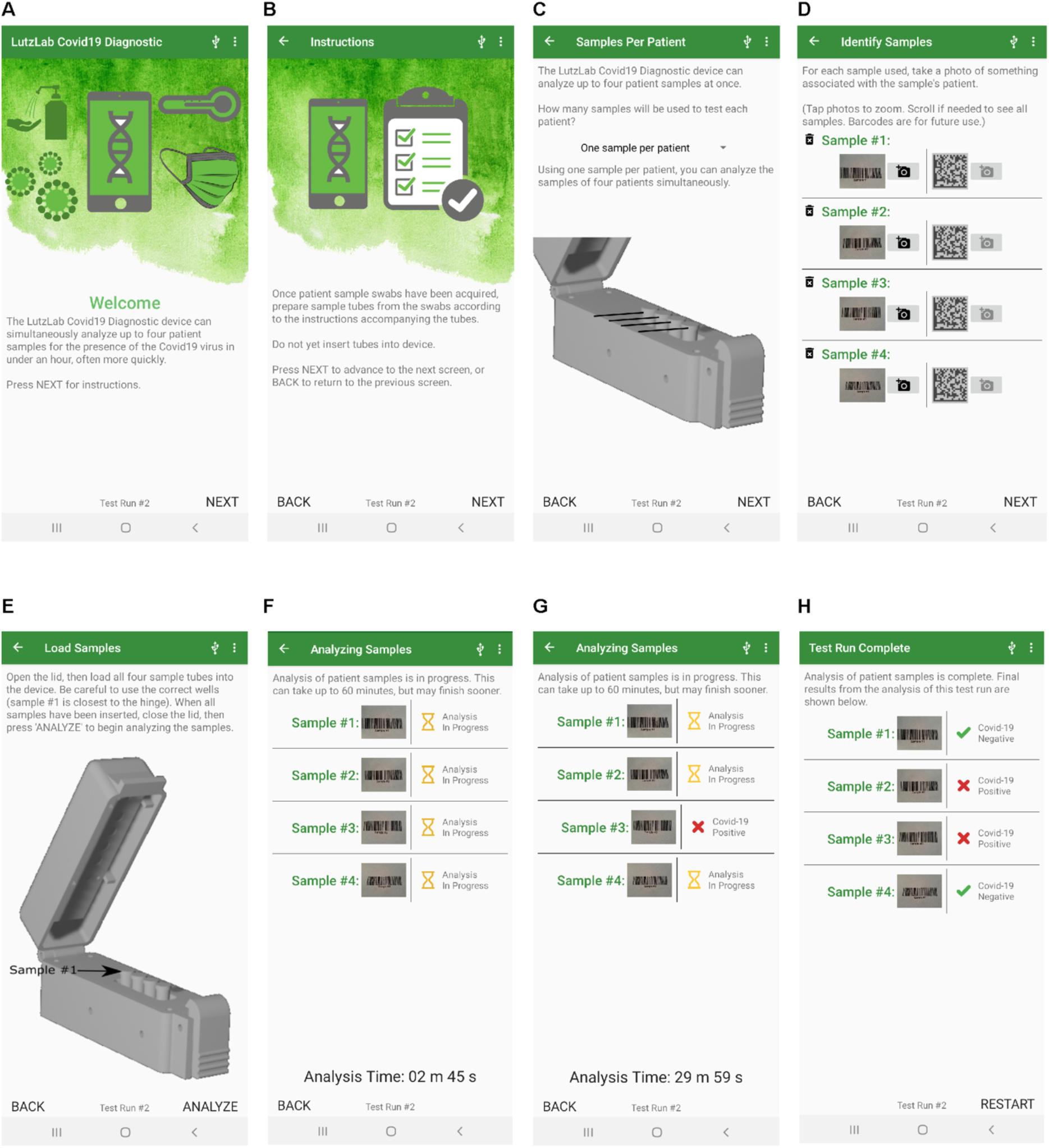
Cell phone user interface and result reporting. The Android Harmony application guides the user through steps to set up and run the test. (**A**) Intended use of the device. (**B**) Instructions for the user to prepare samples. (B) Selection of the number of samples. (**D**) Collection of sample identifiers by barcode, photo capture, or manual typing. (**E**) Instruction to insert the tube when the temperature of the device reaches the reaction temperature. (**F**) Real-time analysis during the test run. (**G**) Positive results are reported as soon as they are detected. (**H**) Negative and IND results are reported at the end of the run time.

### Evaluation using extracted RNA from clinical specimens

Clinical specimens collected from individuals positive for SARS-CoV-2 (*n = 33*) or for other pathogenic respiratory infections (*n = 68*) were re-tested by RT-qPCR(*9*) to measure the SARS-CoV-2 viral load and provide the reference test result. Using RT-qPCR, 68 specimens were confirmed to be SARS-CoV-2 negative (**Fig. 5A**, left), 30 specimens were confirmed to be SARS-CoV-2 positive (**Fig. 5A**, right), and three previously-positive specimens were only detected by one RT-qPCR assay (either N1 or N2) and classified as inconclusive (**INC**, **Fig. 5A**, middle). Specimens were tested in duplicate on Harmony using the ready-to-use reagents (**Fig. 5A)**, arranged from low to high virus concentration as determined by RT-qPCR. Out of 30 RT-qPCR-positive specimens, Harmony detected 25 specimens in both technical replicates and 4 specimens in one of the two replicates. In the three RT-qPCR INC results, one sample was detected by Harmony in one of the two replicates. Excluding the RT-qPCR INC specimens and applying a stringent requirement for both Harmony replicates to be detected to report a positive test, Harmony achieved 95% accuracy (93/98, 95%CI: 88-98%), 83% sensitivity (25/30, 95%CI: 65-94%), and 100% specificity (68/68, 95%CI: 94-100%). If replicates are treated independently, Harmony detected 54/60 positive specimens (90% sensitivity, 95%CI: 79-96%). For specimens with ≥10 copies/rxn, Harmony detected 24/25 specimens (96% sensitivity, 95%CI: 80-100%), and for specimens with ≥40 copies/rxn, Harmony detected 18/18 specimens (100% sensitivity, 95%CI: 81-100%).

**Fig. 5.**
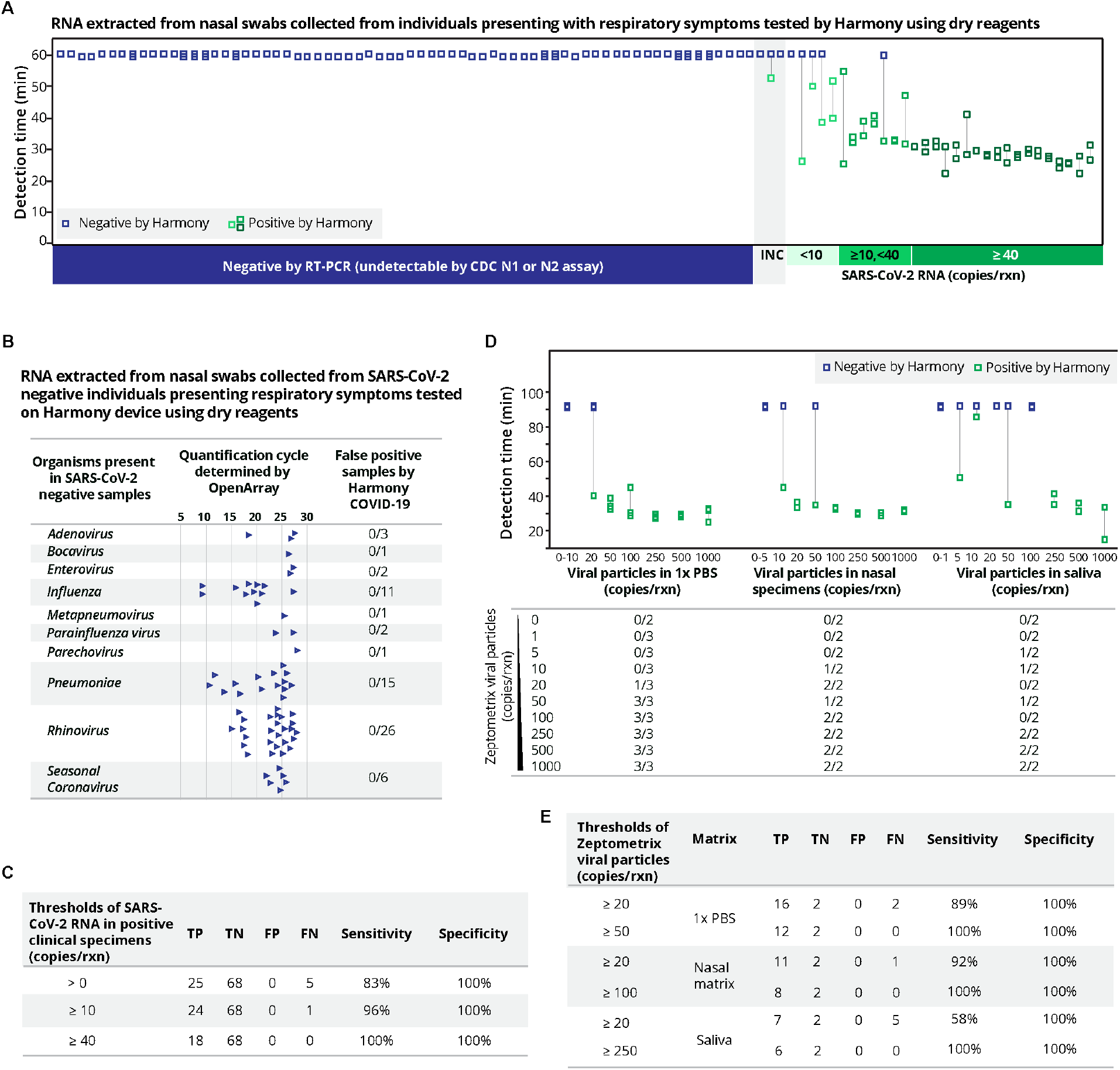
Harmony COVID-19 performance on clinical and blinded contrived specimens. (**A**) Detection time called by Harmony COVID-19 software for the analysis of purified RNA extracted from human nasal specimens from 101 patients presenting respiratory symptoms. Stored specimens were re-tested by RT-qPCR using N1 and N2 CDC primers; results matched previous results in 30 previous positive specimens (positive for N1 and N2) and 68 previous negative specimens, with three previous positive specimens classified as inconclusive (INC) because they were negative for either CDC N1 or CDC N2 by RT-qPCR. Samples are ranked from left to right by increasing SARS-CoV-2 concentrations as determined by RT-qPCR. Extracted RNA from specimens was analyzed by Harmony COVID-19 (10µL RNA in 40µL RT-LAMP dry reagents, *n = 2*). Samples were classified as positive by Harmony only if both replicates were detected. The classification results by Harmony were compared to RT-qPCR using CDC primers and probes (5µL RNA in 20µL RT-qPCR reaction). (**B**) Organisms in clinical samples collected from individuals presenting with respiratory symptoms were quantified using OpenArray, as previously described(*16*). High quantification cycle values indicate low concentrations. All these specimens were correctly identified as SARS-CoV-2 negative by Harmony. (**C**) Summary table for the sensitivity and specificity of the Harmony COVID-19 system using extracted RNA from clinical specimens (RT-qPCR INC samples excluded). (**D**) Blinded analysis of XPRIZE contrived sample panel. The assay was run for 90 min (30 min longer than **Fig. 5A** to accommodate inhibitors that may be present in these blinded samples). The table below shows the detectable samples over the total number of tested technical replicates. (**E**) Summary table for the sensitivity and specificity of the Harmony COVID-19 system on the XPRIZE contrived samples.

The clinical panel (**Fig. 5A**) included 68 specimens that were SARS-CoV-2 negative but contained other respiratory pathogens including influenza, rhinovirus, and seasonal coronavirus (**Fig. 5B**). Harmony correctly identified 68/68 specimens (**Fig. 5A**, left) as negative (100%, 95%CI: 94-100% specificity). **Fig. 5C** summarizes the performance of the extracted clinical specimens.

### Evaluation of contrived SARS-CoV-2 viral particle specimens without extraction

A panel of 69 contrived samples from the COVID-19 XPRIZE competition were tested on the Harmony system with ready-to-use reagents (**Fig. 5, D** and **E**). For SARS-CoV-2 virus in PBS, Harmony detected 100% of samples containing ≥50 SARS-CoV-2 viral particles/rxn, and 89% of samples containing ≥20 viral particles/rxn in PBS (despite PBS being a non-ideal buffer). In nasal matrix, Harmony COVID-19 detected 100% of samples containing ≥100 viral particles/rxn, and 92% of samples containing ≥20 viral particles/rxn. In human saliva, Harmony detected 100% of samples with ≥250 viral particles/rxn and 58% with ≥20 viral particles/rxn. Harmony had zero false positives across all samples.

### Feasibility testing: sample-to-result system tested by healthcare workers and laboratory personnel

To complete the system, we developed a test kit that included all components needed for running a test, including the swab, elution buffer, a fluid transfer device, and a ready-to-use reagent tube. Swabs were pre-loaded with SARS-CoV-2 DNA to serve as a control sample to evaluate performance by HCWs and laboratory personnel (**LPs**).

In the first phase of user testing (**Fig. 6A**), we compared the accuracy and reproducibility of two methods for transferring the eluate to the reaction tube when operated by LPs *(n = 5*) and HCWs (such as nurses, medical students, and dental students; *n = 10*). The first method (**Fig. 6B** and shown-previously in **Fig. 1**) used a unified sampler-dispenser. The second method (**Fig. 6C**) used a transfer pipette to transfer a fixed volume of fluid to the reaction tube. Regardless of the baseline skill of operators, transfer pipettes yielded more reproducible recovered volumes than the unified dispenser (F-test, p <0.001). Recovered volumes of the two methods were not significantly different between HCWs and LPs (student’s t-test, two-sided, p = 0.90; **Fig. 6D**). User feedback on the transfer methods and preferences on tube sizes are summarized in **table S2**.

**Fig. 6.**
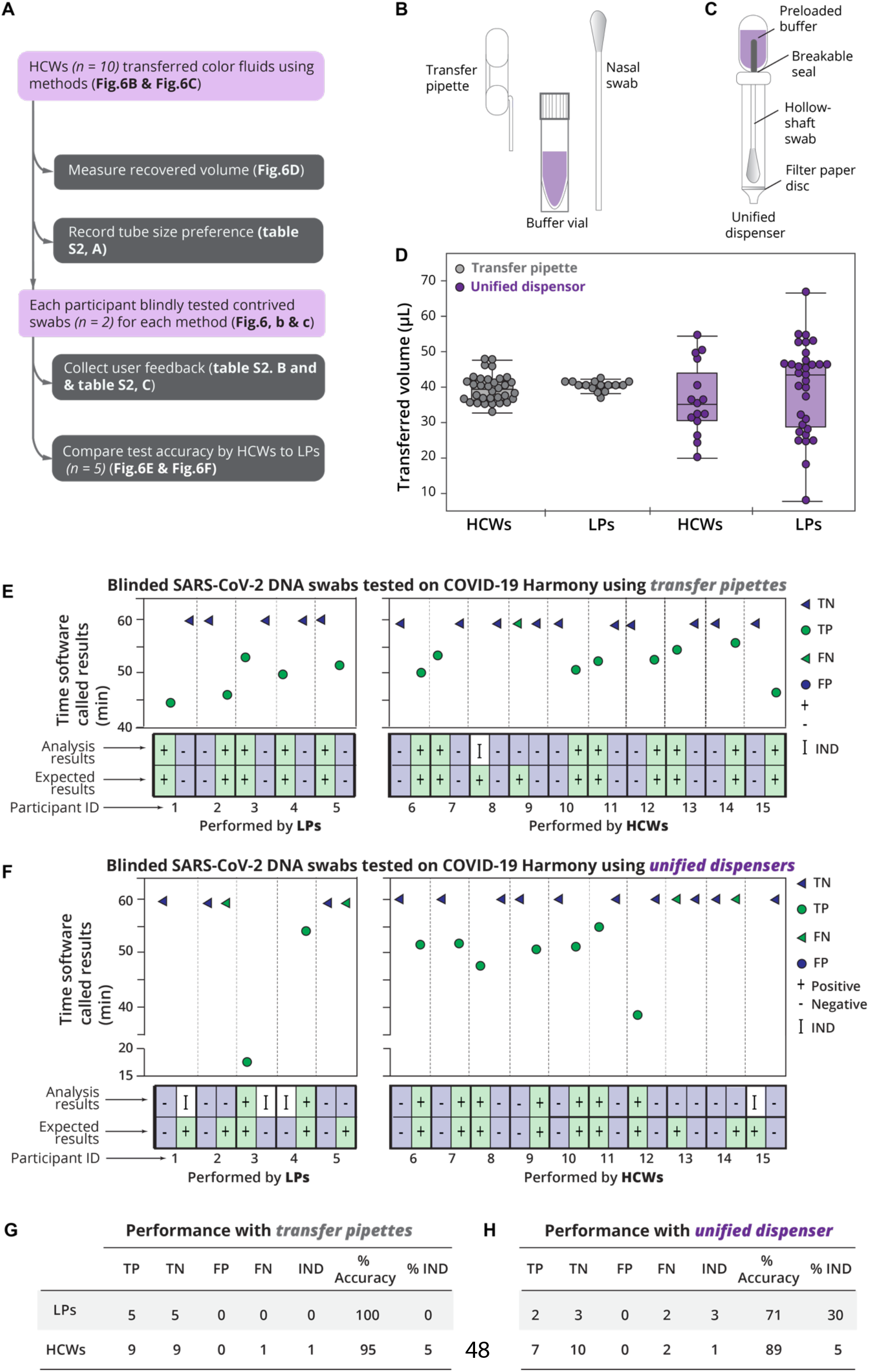
Usability study of Harmony COVID-19 workflow including two sample transfer methods at the POC. (**A**) Study design. (**B**) Illustration of the first sample transfer method including a volumetric transfer pipette (40µL), a nasal swab, and a pre-loaded buffer vial. **video S1** demonstrates this workflow. Briefly, swabs are manually rubbed on the side of the tube for 30 seconds, soaked for 1 minute, and removed. The pipette is then used to transfer the swab eluate to rehydrate the lyophilized RT-LAMP reaction tube. (**C**) Illustration of the second sample transfer method option using a unified dispenser, which integrates a swab, buffer container, and a dropper in a single device. The contained buffer is released to elute the swab sample by cracking a conduit, the user agitates the fluid by shaking the tube, and a dropper tip allows the use to transfer two drops of fluid to the reaction tube by squeezing the body of the device. **video S2** demonstrates this workflow. (**D**) Volume transferred by each method performed by healthcare workers (**HCWs**) or laboratory personnel (**LPs**). Individual volume measurements are plotted along with medians (middle lines of the boxes), interquartile ranges (edges of the boxes), and ranges (the ends of whiskers). **e** and **f**, Assay performance on blinded contrived swabs (one negative swab and one positive swab with SARS-CoV-2 DNA at 1,000 copies/swab, corresponding to 100 copies/rxn) by LPs (*n = 5*) and untrained HCWs (*n = 10*) using transfer pipettes, (**E**) or unified dispensers, (**F**) The usability experiment written protocol is available on protocol.io (https://dx.doi.org/10.17504/protocols.io.bkvskw6e) and as a visual demonstration in **video S1 and video S2**. (**G**) Performance summary for the transfer pipette method. (**H**) Performance summary for the unified dispenser method.

Next, HCWs and LPs executed the full Harmony workflow on contrived SARS-CoV-2 DNA swab samples (**Fig. 6**). Using the transfer pipette workflow (**Fig. 6E**), the test accuracy was 100% (10/10) for LPs and 95% (19/20) for HCWs. Surprisingly, we found that the HCWs performed better than the LPs when using the unified dispenser system (**Fig. 6F**), both accuracy and IND rates. HCWs had significantly lower (Z-test, two-tailed, p<0.05) IND results (1/20) than LPs (3/10) when using the unified dispenser system. Excluding IND results, the HCWs had 89% accuracy, higher than LPs with 71% accuracy (Z-test, two-tailed, p <0.05), when using the unified dispenser system.

## DISCUSSION

This work presents a combination of multi-disciplinary engineering (molecular, mechanical, electrical, software, human interface design) to develop a complete system for SARS-CoV-2 detection at the POC. The system is inexpensive and simple yet has features comparable to laboratory-based tests (**Table 1A**). The assay has high sensitivity and specificity in clinical and contrived specimens, and HCWs successfully performed the workflow with high accuracy.

**table 1.**
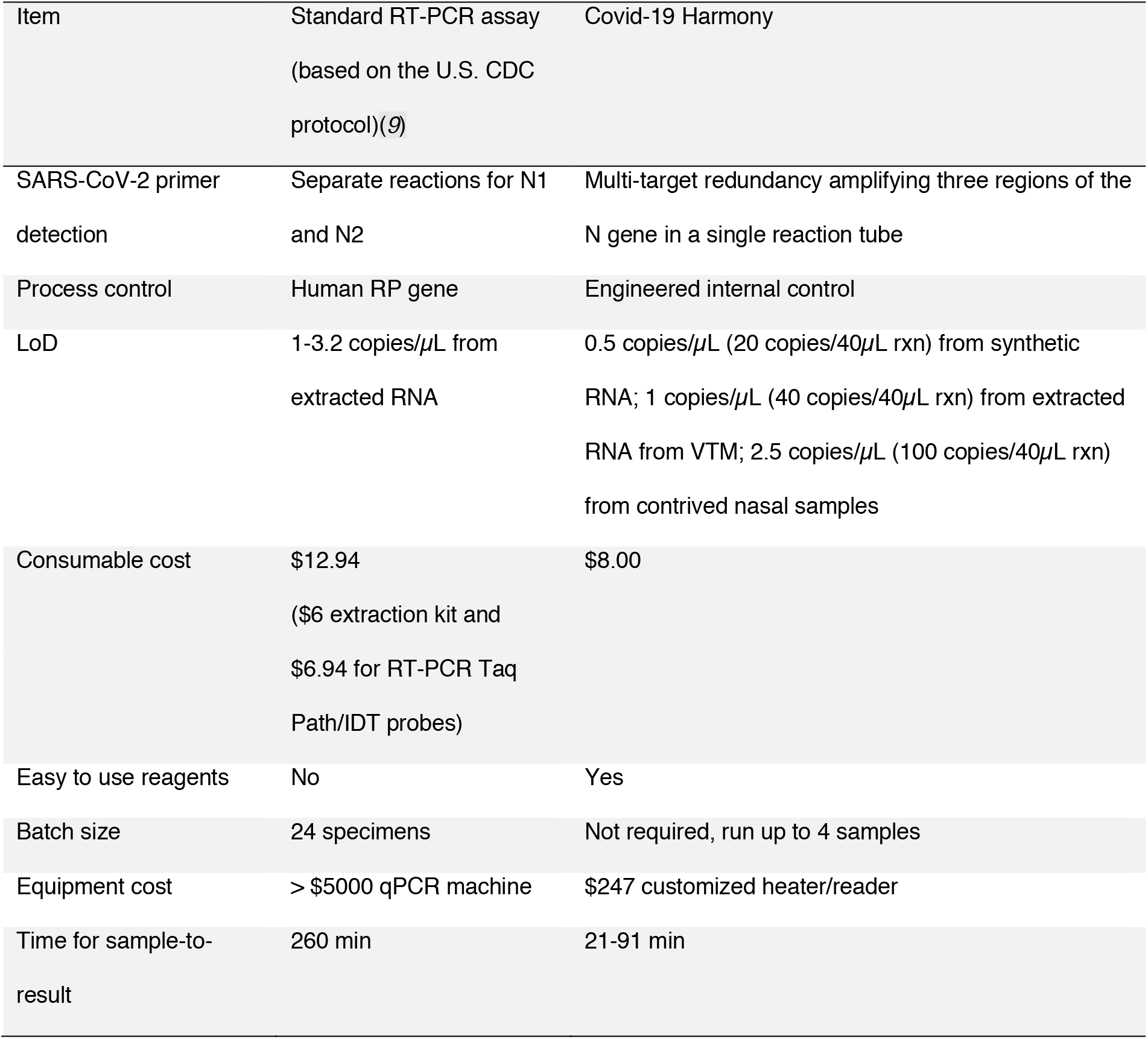
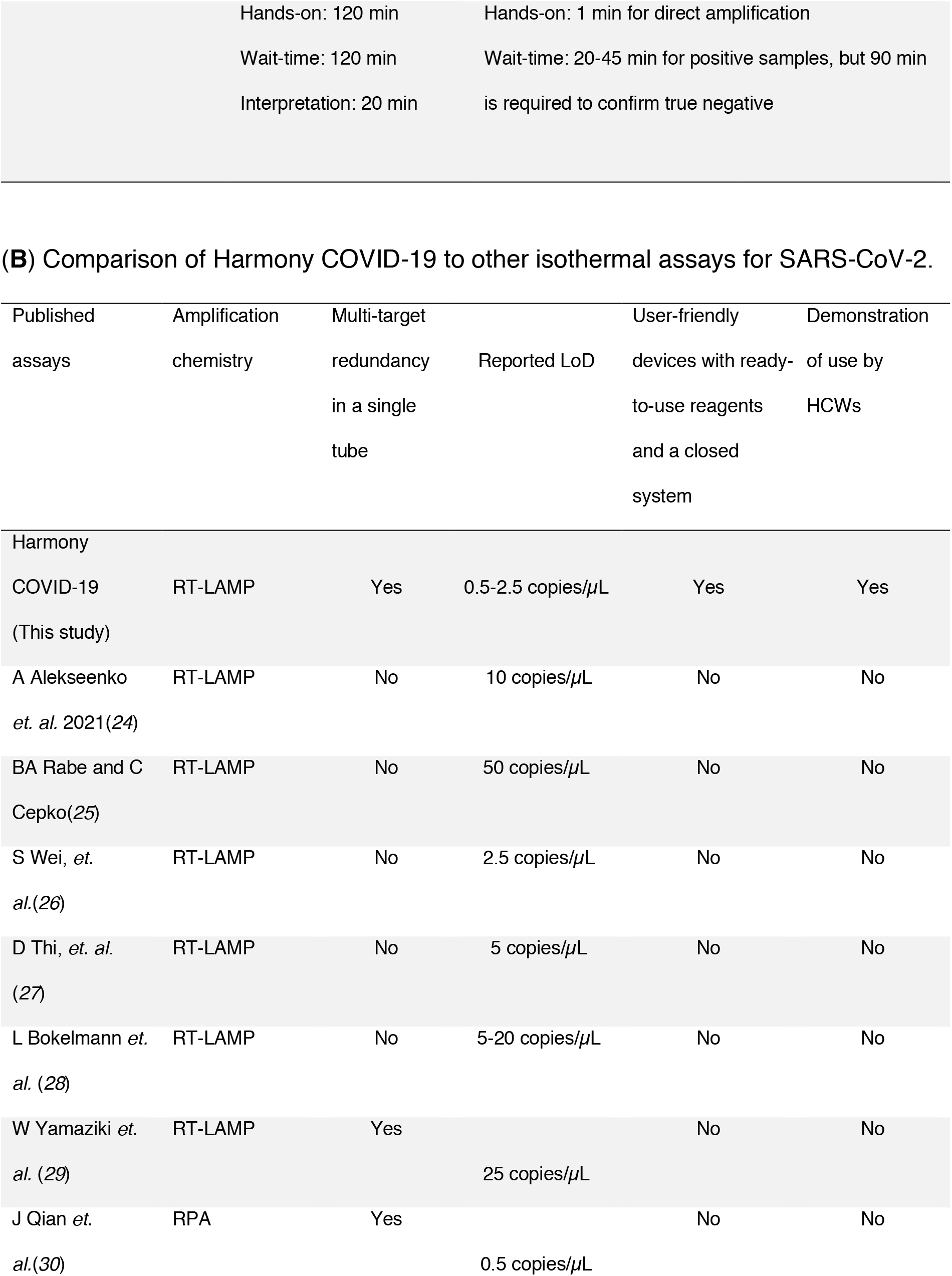
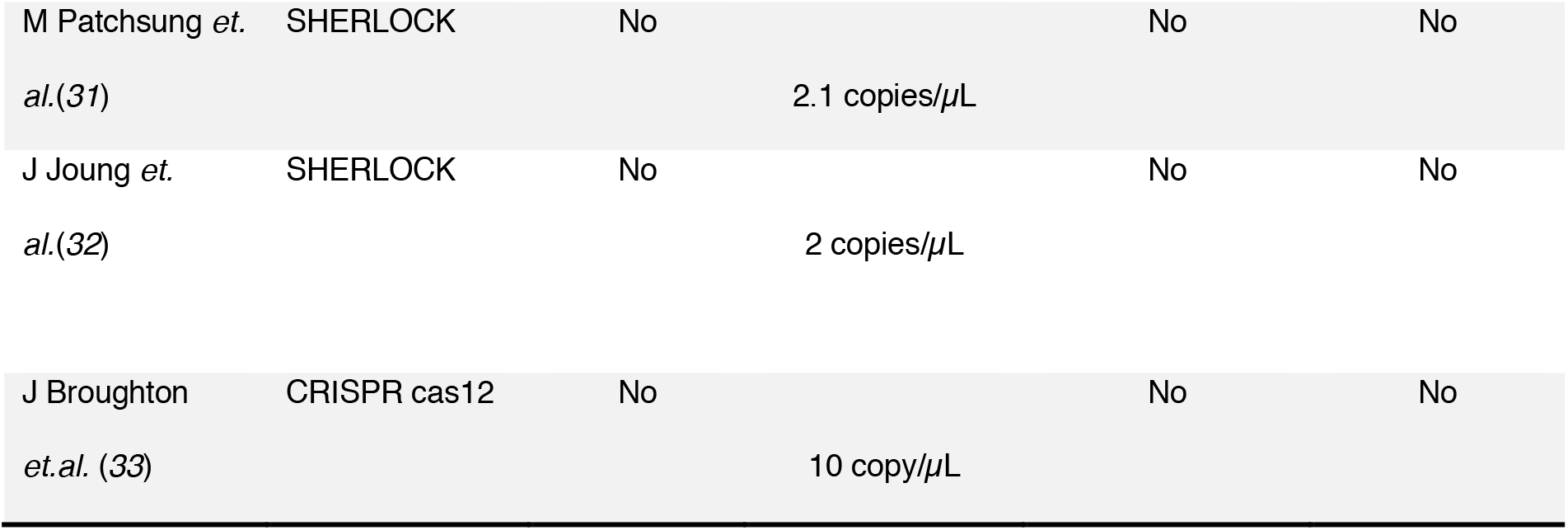
Feature comparison between the Harmony COVID-19 system and other assays. (**A**) Comparison to the U.S. CDC assay.(*9*) Full cost calculations for reagents and devices are enclosed in **tables S3** and **table S4**.

Harmony uses ready-to-use reagents that improve speed and accuracy for assay setup. Unlike other lyophilized RT-LAMP^25^, our lyophilized reagents are magnesium-free, which prohibits any enzymatic activity that may occur before assay initiation. Our lyophilized RT-LAMP is activated upon the addition of sample which contains magnesium and detergent. Thus, it should be less prone to moisture degradation than magnesium-containing lyophilized RT-LAMP.

The lyophilized Harmony assay detected as low as 15 copies/rxn of synthetic SARS-CoV-2 RNA and detected all extracted RNA samples from clinical specimens at ≥40 copies/rxn. Here, we used only 10µL RNA in 40µL reaction to allow head-to-head sensitivity comparison to the CDC RT-PCR protocol that uses 5µL RNA in a 20µL reaction. The lyophilized reagents can accommodate up to 40µL of sample (4x than used here), which could further increase Harmony sensitivity.

Harmony lyophilized RT-LAMP is robust against simulated nasal matrix containing mucin, salt, and human genomic DNA. The impact of salts and mucin was observed as a delayed reaction time for both target and IAC. To account for this delay, we adjusted the assay time to 90 min when analyzing specimens with nasal matrix. We found excellent sensitivity when testing samples with ≥50 viral particles in nasal matrix (estimated 1250 copies/swab for 1mL elution buffer and 40µL reaction), which would be sufficient to detect SARS-CoV-2 in most nasal specimens (10^3^-10^9^ SARS-CoV-2 copies/swab) from individuals within the first 3-5 days after the symptom onset^26^. Further, Harmony detected all samples with ≥100 viral particles in nasal matrix (estimated 2500 copies/swab for 1mL elution buffer and 40µL reaction). While saliva was not our intended specimen type, we found a moderate sensitivity when testing the XPRIZE blinded saliva panel. Heat treatment of saliva shown to enable detection by RT-PCR^27^ would likely improve the sensitivity of our RT-LAMP in detecting SARS-CoV-2 in saliva.

Isothermal amplification, like RT-LAMP in Harmony, is often promoted as ideal for POC testing because it can be carried out using a constant temperature heat source, such as a water bath, heat block, or even more novel sources (chemical heat source, sunlight, body temperature)(*10*). However, reported tests often have other limitations that may prevent their use in POC testing (**Table 1B**). Common limitations include use of reagents use of refrigerated or frozen reagent stocks that must be formulated using laboratory pipettes, manual steps for sample processing that require laboratory skill, or detection steps that rely heavily on user interpretation or analysis by a user’s cell phone. Harmony is a complete sample-to-result system that uses ready-to-use reagents for simple 1-minute setup and a dedicated device that reports results without extra post-amplification steps or reliance on user interpretation.

Some RT-LAMP tests use end-point lateral flow strips for detection^28^, which adds a user step, but more importantly, should not be done at the POC because it exposes the testing site to amplicons that will give false positive results in subsequent tests. Others use in-tube detection by eye(*11*),(*11*) or using a cell phone(*12*) which increases the user burden and could introduce additional sources of error. Real-time detection used in Harmony COVID-19 removes the need for extra detection steps or user interpretation.

Real-time detection also allows Harmony to report positive results as soon as the SARS-CoV-2 signal appears rather than waiting for an end-point detection method. High viral load samples could be detected in as little as 20 min, which could enable more timely infection control measures to limit viral spread. Harmony real-time detection allows testing without opening tubes after amplification, requires no extra steps or interpretation by the user, and allows reporting a positive sample earlier than end-point detection methods.

The Harmony device automates heating, multiplexed detection, and data interpretation. We integrated a simple heater with low-cost LEDs and sensors to enable real-time two-color detection of RT-LAMP fluorescence in four independent sample wells. The device costs <US$300 in parts for prototypes, while the lowest cost commercial RT-PCR machine on the market is >US$5000(*13*). Moreover, the Harmony device is portable and energy-efficient, requiring <15 watts and operable by USB power sources. These features are attractive for use at the POC in the U.S. and resource-limited settings.

Lastly, the simple workflow and analysis of Harmony is appropriate for POC testing. Test operation involves a few simple steps that take ∼1 minute, and there are no further user steps after starting the test. Here, HCWs were able to operate the tests without training using only simple written instructions and watching the procedure on video. The simplicity of the Harmony procedure and completeness of the stand-alone system can be a significant advantage for onboarding to ambulatory care settings and potentially community testing locations.

In this study, we evaluated the system using RNA extracted from clinical samples stored in viral transport medium (**VTM**) and contrived specimens in unknown nasal and saliva matrix (XPRIZE), which may not reflect performance in fresh clinical samples. These samples were used due to the difficulty of obtaining fresh dry swabs intended for this POC test, and RNA was extracted from clinical samples because VTM is irrelevant for immediate testing at the POC. Nevertheless, this testing shows excellent analytical reactivity to clinical specimens, lack of cross-reactivity with other respiratory pathogens, analytical sensitivity comparable to the CDC RT-PCR, and resilience in the presence of nasal matrix.

While our assay is relatively fast and sensitive, there is potential to improve performance. We ran RT-LAMP at 64°C, which is 9°C above the optimal temperature for the RT enzyme. We hypothesize that cooling of the heat block when the lid is open, followed by a few minutes to ramp back up to 64°C, provided an opportunity for the RT enzyme to operate. In contrast, the reaction speed of the temperature-stable DNA polymerase in this assay is optimal at higher temperature. Thus, operating the device with an initial 55°C step for optimal RT activity followed by a higher temperature step for fast DNA amplification could increase sensitivity and/or speed.

The current design of RT-LAMP assay can be improved to include an IAC that is more representative of the SARS-CoV-2 virus. We currently use a DNA IAC, which has been used in several EUA assays^34^, but the DNA control does not capture the potential failure of the lysis and cDNA conversion processes. We are developing a new IAC that uses RNA that can be packaged in a viral envelope. This modification may require re-development of excipients that preserve the RT-LAMP enzymes and the enveloped RNA.

POC tests like Harmony could provide rapid testing to enable the re-opening of businesses, schools, and borders; but the need for POC testing extends beyond the current emergency. As natural immunity and vaccination take hold, COVID-19 is likely to become endemic with seasonal outbreaks. Tests such as Harmony will be necessary not only to help identify individuals with SARS-CoV-2, but other respiratory pathogens such as influenza and RSV, as the symptoms of these are similar. Prior to the current SARS-CoV-2 pandemic, the yearly influenza infections caused nearly 30 million cases in the U.S. alone, with 400,000 hospital admission and 35,000 deaths(*14, 15*). Indeed, particularly when more typical patterns of respiratory infections occur, there will be a need for tests that can perform multiplex testing for individuals presenting respiratory symptoms. Tests such as Harmony will become a common practice for confirming presence of viral infections that can be treated with antiviral therapies or that demand infection control, as well as helping to reduce inappropriate use of antibiotics. Further, test platforms developed for this pandemic could be more readily adapted to detect new diseases to increase readiness for future pandemics. The advances in awareness and acceptance of testing for infections at scale, and the technology platforms developed will have ongoing benefit for COVID-19 control, reducing harm from other endemic diseases, and fighting future pandemics.

## MATERIALS AND METHODS

### Preparation of synthetic RNA standards

MERS and SARS-CoV-1 plasmid standards (10006623 and 10006624, Integrated DNA Technologies, Coralville, IA) were amplified using M13 PCR primers to generate DNA templates tagged with T7 promoter sequences. DNA templates containing T7 promoter sequences were transcribed using Hi-T7 RNA polymerase (M0658, New England Biolabs, Lawley, MA) following the manufacturer’s protocol, with 1 U/*µ*L RNase inhibitor (N2611, Promega, Madison, WI). Following transcription, 0.01U/*µ*L DNase I (EN0521, ThermoFisher Scientific, Waltham, MA) was added to each reaction, and incubated at 37°C for 15 min. RNA transcripts were purified with Monarch RNA Cleanup Kits (T2040, New England Biolabs). RNA was quantified on a Qubit 4 fluorometer (Q32852, RNA HS Assay Kit, ThermoFisher Scientific, Waltham, MA), and checked for length and integrity by electrophoresis on TapeStation (5067 RNA screening tape, Agilent Technologies, Santa Clara, CA). RNA at 10^10^ copies/*µ*L was stored in nuclease-free water in single-use aliquots. SARS-CoV-2 RNA was prepared and quantified as previously described(*16*).

### In-house polymerase production

#### Plasmid preparation

The chimeric polymerase (TFv1) was generated by assembly PCR using genomic material from *Thermus thermophilus* HB27 (ATCC BAA-163D-5™, Gaithersburg, MD) and a synthetic gene fragment derived from *Thermodesulfatator indicus* (*T. in*), (Integrated DNA Technologies, Coralville, IA), similar to the method previously described(*17*). Briefly, a 3’ terminal fragment of the *T.th* HB27 ***Pol*A** gene downstream of the finger domain was amplified by PCR using Phusion® Hot Start Flex DNA Polymerase (M0535L, New England Biolabs) with a modified forward primer containing sequence overlap with the *T. in polA* gene fragment *(AA734-795,* codon optimized for *E. coli)*. A chimeric sequence was then generated by assembly PCR. A second*T.th* polymerase fragment, upstream of the finger domain and lacking the 3′to 5′ exonuclease domain, was similarly amplified and appended to the 5’ end of the chimeric fragment. The resultant sequence corresponds to a *T. th* DNA polymerase I fragment, orthologous to the Stoffel fragment of *T.aq.* polymerase, with a replacement in the finger domain derived from *T. in*. This gene was then cloned into the expression vector pET His6 TEV LIC cloning vector (2B-T) (a gift from Scott Gradia, Plasmid # 29666, Addgene, Watertown, MA) by ligation independent cloning(*18*). The annealed vector and insert were transformed into T7 Express lysY/Iq Competent *E.* coli (C2987H, New England Biolabs) per the manufacturer’s protocol and plated on LB 100µg/mL carbenicillin plates. Colony plasmid inserts were verified by Sanger sequencing.

#### Protein expression and purification

50mL starter culture in LB broth with 100µg/mL carbenicillin was inoculated with a sequence-verified colony and incubated at 30°C overnight in a shaking incubator at 200rpm. The culture was centrifuged at 5,000 RCF for 15 min and resuspended in an equivalent volume fresh LB carbenicillin media. Flasks of 500mL LB broth with 100µg/mL carbenicillin were inoculated with 5mL of resuspended starter culture and incubated at 37°C in a shaking incubator at 200rpm until OD_600_ of 0.4-0.6 had been achieved. Cultures were induced with 0.4mg/mL of IPTG and incubated for two hours then harvested by centrifugation at 5,000 RCF for 15 min, decanted, and stored as cell pellets at –80°C until needed. Prior to purification, cell pellets were resuspended in lysis buffer (20mM sodium phosphate pH7.4, 80mM NaCI, 1%*(v/v)* Triton (648466 Sigma, St. Louis, MO), and 1mg/mL lysozyme from chicken egg white (Millipore Sigma, Burlington, MA)). Suspensions were heated at 37°C for 20 min, then lysed by ultrasonic disruption using a Fisherbrand™ Model 120 Sonic Dismembrator (Fisher Scientific, Waltham, MA) with two cycles of two second pulses at 50% amplitude for 2 min, while on ice. Lysates were heat-clarified at 65°C for 20 min, followed by centrifugation at 20,000 RCF. The supernatant was collected and diluted 1:6 with sample diluent buffer (20mM sodium phosphate, 1.8M NaCI, pH 7.4) and filtered with a 0.22 µm syringe filter (25-342 Olympus, Genesee Scientific, San Diego, CA). The Purified clarified lysate was applied to a 1mL Hitrap Talon Crude column (Cytiva, Marlborough, MA) and enriched per manufacturers default recommended specifications using an ÄKTA start system with the Frac30 fraction collector (Cytiva). Elution fractions were evaluated by A_280_ on a spectrophotometer (2200 Nanodrop, Fisher Scientific) using the proteins expected molecular weight (63.062kDa) and extinction coefficient (60,850m^2^/mol) of the His-tagged polymerase. Elution fractions containing protein were diluted 1:10 with 10mM sodium phosphate and applied to a 1mL HiTrap^®^ Heparin HP column and purified per the manufacturers recommended specifications except for a modified binding buffer (10mM sodium phosphate, and 30mM NaCI, at pH 7). Fractions containing protein were concentrated and buffer exchanged into storage buffer (17mM Tris-HCI, 167mM KCI, 17% glycerol pH7.5) using an Amicon® Ultra-2 30K device (Millipore Sigma), as recommended by the manufacturer. The recovered polymerase solution was then adjusted to 10mM Tris-HCI, 100mM KCI, 2mM DTT, 0.1%Triton X-100, 50%glycerol pH7.5. Polymerase purity was evaluated by SDS-PAGE and spectrophotometry throughout purification.

### RT-LAMP assay design

Alignment of SARS-CoV-2 sequences (available on NCBI up to January 2020) were performed using Geneious (Auckland, New Zealand). Three sets of LAMP primers (Harmony NC1, Harmony NC2, and Harmony NC3) were developed targeting the 22,285 – 28,517 (232 bases), 28,528 – 28,471 (213 bases), and 29,135 – 29,416 (281 bases) regions of the SARS-CoV-2 nucleocapsid phosphoprotein gene. *In-slico* analysis showed that this primer design matched 100% of SARS-CoV-2 sequences and did not have cross-reactivity to MERS and SARS-CoV-1. We used two detection probes, each comprised of a fluorescent probe and its complementary quencher probe. For detection of SARS-CoV-2 RNA amplification, we used FAM labeled detection probe and Iowa Black™ quencher probe. For detection of IAC amplification, we used TEX 615 labeled detection probe and BHQ2 labeled quencher probe. The primer and probe sequences are listed in **table S1**.

### Lyophilization of RT-LAMP reagents

Reactions containing 2x primers (3.2mM each FIP/BIP primer, 0.4mM each loop primer, and 0.8mM each F3/B3 primer) and probes (0.4mM fluorophore probes and 0.6mM quencher probes) in the presence of 5% *(v/v)* mannitol (OPS Diagnostics EXMN 500-01), 10mM DTT, 1.4µg DNA polymerase, 1.2U/*µ*L WarmStart RTx (M0380L, New England Biolabs), 2.8mM each dNTP in 10mM Tris-HCl, pH 7.4, 2U/*µ*L RNase inhibitor (N2615, Promega), 50mU/*µ*L thermostable inorganic pyrophosphatase (M0296L, New England Biolabs), and 2500 copies/*µ*L IAC DNA were prepared in 20µL aliquots of 0.2mL reagent tubes and lyophilized. Packages were stored in desiccant packets at - 20°C until use.

### RT-LAMP reaction setup

Lyophilized reagents were resuspended in a final volume of 40µL containing 1x rehydration buffer (ThermoPol buffer (B9004S, New England Biolabs), 6mM MgSO_4_, and 0.5%*(v/v)* Triton X-100). For analytical sensitivity experiments, 2µL SARS-CoV-2 RNA in water was added. For clinical testing, 10µL extracted RNA was added to each

RT-LAMP reaction. For testing the contrived specimens, 10µL sample (not extracted) was added to each RT-LAMP reaction. For usability testing, 40µL swab eluate was added directly to the RT-LAMP reaction using transfer pipettes or the in-house unified dispensers.

### Analytical sensitivity of RT-LAMP assay

Analytical sensitivity of lyophilized RT-LAMP was tested using purified RNA (0-2x10^6^ copies/40µL reaction). We also tested the effect of salt, human DNA, and mucin on amplification of SARS-CoV-2 RNA and IAC DNA using simulated nasal matrix as previously described(*19*).

### Preparation of contrived swab samples and kits for usability testing

For the transfer pipette workflow, each kit contained one swab (either positive or negative), a vial of buffer (10-500-25, Fisher Scientific, Waltham, MA) containing 400µL of 1x rehydration buffer (see section RT-LAMP reaction setup) and one lyophilized RT-LAMP reagent tube. For positive swabs, 10µL of 100 copies/*µ*L SARS-CoV-2 DNA in 0.05% Triton X-100 was pipetted onto the swab (1804-PF, Puritan Medical) and dried at room temperature in the air-clean chamber (825-PCR/HEPA, Plas-Labs, Lansing, NI) for 4 hours before packaging in a sealed foil pouch with desiccant (S-8032, ULINE). The unified sampler was built from parts using an assembly jig (custom order from Hygiena, LLC, Camarillo, CA). The unified dispenser contained a hollow-shaft polyurethane swab attached to a bulb preloaded with 400µL 1x rehydration buffer and a separate tube with a filter and drip dispenser friction fit inside the tube. Each custom sampler kit contained an in-house assembled dispenser unit and one lyophilized RT-LAMP reagent tube.

### Usability study

The Harmony usability study was approved by the Institutional Review Board at the University of Washington (IRB#: STUDY00010884), and informed consent was provided by participants. HCWs (*n = 10*), including registered nurses, medical students, and dental students were enrolled in this study over the course of 1 month. All data was collected without identifiers. The primary objective of this study was to determine the accuracy and precision of the two sample transfer methods and test if variation in reproducibility impacted assay performance, and the secondary objective was to gather user input on the feasibility of each protocol. Participants first received an explanation of the test and its intended use, then watched an instructional video narrated by the facilitator to become familiar with the Harmony workflow (**video S1 and video S2**). After watching the video, participants were given a packet with a unique identifier that contained comprehensive instructions for the study procedures and survey questions. Study procedures were modeled on a coaching methodology(*20*), to allow natural progression of the procedures and to identify blocking steps in the instructions. The facilitator sat 10 feet from the participant and kept a record of verbal questions and comments from participants but did not provide answers to procedural questions unless the participant was could not proceed.

For part 1, data related to user preference on sample preparation, tube sizes, and their level of confidence in performing each task were collected anonymously as prompts within their study packet. Open-ended questions were extracted for themes in errors or issues and supplemented with comments and observations recorded by the facilitator. Samples were saved with deidentified labels, and after all participants had completed the study facilitators measured volumes in each receiving tube from part 1. The variances of the volume recovered by each method were tested for their equality using F-Test, and the P-value was reported. The means of the recovered volumes transferred by LPs or HCWs were compared using student’s t-test (two-sided), and the P-value was reported.

For part 2, each participant followed instructions in the packet and on the integrated mobile phone to complete the entire Harmony workflow on four total contrived samples, two samples using the transfer pipette protocol and two using the unified sampler system. Kits were prepared as specified above.

The lab personnel (*n = 5*) portion of the user testing aimed to compare performance of the Harmony device by HCW to that of an “experienced” population, by replicating this process with four students and a researcher from the Lutz Lab. Survey questions were omitted for this group because these volunteers provided feedback throughout the development of Harmony and their views are included in the discussion of this manuscript. The accuracies of tests performed by LPs and HCWs using each method were tested for their equality using Z-test (two-sided), and the P-value was reported.

### Clinical nasal swab specimens from individuals presenting respiratory symptoms

A minimum sample size of 30 (each negative and positive) was required to achieve the confidence interval (**CI**) of 90 -100% (binomial cumulative distribution function) should all results be accurate. Our clinical specimen panel (*n = 110*) was previously used to evaluate other SARS-CoV-2 assays(*16*), but 9 specimens had insufficient volume for RNA extraction and were thus excluded from the study. These specimens were collected with informed consent as part of the Seattle Flu Study, approved by the Institutional Review Board at the University of Washington (IRB#: STUDY0006181). All participant specimens were de-identified prior to the transfer to the Lutz lab at the University of Washington for analysis. The remaining 101 specimens were evaluated using the newly-extracted RNA for analysis by RT-qPCR and Harmony system, to avoid variations introduced from different batches of extraction. RNA was extracted from clinical nasal swab specimens stored in viral transport media using the QIAamp Viral RNA Mini Kit (52906; Qiagen, Hilden, Germany). Prior to extraction, 100µL of each specimen was mixed with 40µL negative VTM to reach the suggested 140µL sample volume. RNA was extracted according to the manufacturer’s protocol, eluted into 70µL of EB buffer, and stored at -80°C in single-use aliquots until amplification. 5µL of RNA was used for analysis in 20µL RT-qPCR using U.S. CDC N1, N2, or RP human control assays(*9*). 10µL of RNA was used for analysis in 40µL lyophilized RT-LAMP in the Harmony workflow. Each sample was run in duplicate. Only samples with positive results from both Harmony replicates were reported as positive for SARS-CoV-2. Sensitivity (TP/(TP + FN)), specificity (TN/(TN+FP)), and concordance ((TP+TN)/(TP+TN+FP+FN)) were calculated and reported with 95%CI using binomial exact proportions.

### XPRIZE blinded contrived sample panel

The blinded panel was assembled and distributed by HudsonAlpha Discovery (Huntsville, AL) for COVID-19 XPRIZE competition, and information on the samples was revealed to the authors only after results were submitted to XPRIZE. Samples were shipped to the Lutz Lab at the University of Washington. We reported results for contrived samples of chemically-inactivated SARS-CoV-2 viral particle (ZeptoMetrix Corporation, Buffalo, NY) spiked in phosphate-buffered saline (**1x PBS**) (*n = 29*), nasal specimens (*n = 20*), or saliva (*n = 20*) at 0 to 1000 copies/rxn. To affirm the identify of these specimens, we also performed RT-qPCR using CDC primer/probes after the samples had been tested using the Harmony system.

### Housing

The housing combines the heater/reader assembly with the cell phone, while concealing the connector cables and presenting a sturdy and easy-to-us0e platform. The 3D CAD program, Fusion 360 (AutoDesk), was used for the design of housing. Prototypes were printed in PLA plastic (Overture™ Filament) using a Prusa i3 MK3s at 0.3mm layer height. Power was supplied to both the heater/reader assembly and the phone simultaneously using the Charge-Plus USB-C (LAVA Computer Manufacturing Inc.) connector device and a standard fast-charging American cellphone charger. The LAVA device also facilitates the transfer of data between the devices.

### Reader/heater device

The aluminum block to house the reagent tubes was custom-made by Bryan Willman according to the specifications in **fig. S3**. The circuit boards were designed using Autodesk Eagle (San Rafael, CA), and the assembly of the circuit boards are shown in **fig. S4** and **fig. S5**. The lid, latch, and upper and lower housing of the device were drawn using SolidWorks (Dassault Systèmes, Waltham, MA) and printed by Xometry - HP MultiJet Fusion 3D (Gaithersburg, MD). All parts were assembled as shown in **fig. S6**.

### Real-time fluorescence signal analysis

After the user inserts the reaction tube into the device, the software adjusts the temperature to re-equilibrate the temperature of heat block back to the reaction temperature. During this initial period, the signals from the photo diodes (the blue LED (FAM signal for target amplification) and the yellow LED (TEX 615 signal for the IAC amplification) are collected but not used in analyses. Once the initial period is over, signal analysis commences. Each signal is analyzed in real time by comparing the current mean of the signal over the most recent 60s interval to the signal’s behavior some 360s in the past. Specifically, the current mean is compared to a bound consisting of the signal’s mean over the period from 420s to 360s in the past plus a multiple (1.9x for target, 1.5x for IAC) of the signal’s sample standard deviation over that same period. If the current mean exceeds the bound, and maintains that condition for at least 15s, the signal is said to have "indicated". If the signal from the target emission indicates, the presence of SARS-CoV-2 has been detected, and the software immediately reports “positive COVID-19”. If the signal from the IAC emission indicates, successful amplification of the IAC has been detected. Then, if the target emission does not also indicate before the end of the run, then a “negative COVID-19” result is reported; this delay in reporting negative results provides the maximum opportunity to detect low concentrations of SARS-CoV-2 that may be present in samples. If neither the target emission nor the IAC emission is detected, "test failed” is reported.

## Data Availability

Data and materials availability: All data are presented in the main text and supplementary information.

## ACKNOWLEDGMENTS

We thank Bryan Willman for his support in fabricating customized heat blocks, Jan Gray for his help with firmware development, Tom Blank for sharing the PCB fabrication resources, and Dr. Liz Sanocki for the useful feedback on the software user interface. We thank Shawna Cooper for sharing the graphics and content used in Audere’s mobile app’s flu@home, which influenced the design of the first few screens of the Harmony COVID-19 software. We thank Dr. Paul Yager and Steven Bennett for use of their optical filters during the initial stage of device development. We thank Jason Tauscher at the Washington Nanofabrication Facility for his assistance in cutting the plastic sheet filters. The Washington Nanofabrication Facility is a National Nanotechnology Coordinated Infrastructure (NNCI) site at the University of Washington with partial support from the National Science Foundation via awards NNCI-2025489 and NNCI-1542101. We thank Seattle Flu Study researchers Dr. Jay Shendure and Dr. Jase Gehring for their helpful technical discussion. We thank Dr. Syamal Raychaudhuri, Dr. Frances Chu from Inbios International, and Dr. Gwong-Jen J. Chang for the stimulating discussion. We thank the Seattle Flu Study and the Seattle Coronavirus Assessment Network (SCAN) teams led by Principal Investigators: Helen Y. Chu, MD, MPH, Michael Boeckh, MD, PhD, Janet A. Englund, MD, Michael Famulare, PhD, Barry R. Lutz, PhD, Deborah A. Nickerson, PhD, Mark J. Rieder, PhD, Lea M. Starita, PhD, Mathew Thompson, MD, MPH, DPhil, Jay Shendure, MD, PhD and Trevor Bedford, PhD for providing specimens for testing. **Funding**: This work was supported by the Seattle Flu Study (funded by Gates Ventures) and the National Institutes of Health (R01AI140845; 5R61AI140460-03). We thank the Washington Entrepreneurial Research Evaluation and Commercialization HUB program (WE-REACH, U01 HL152401) and their matching grant partners for partly supporting reagents, supplies, and personnel. I.T.H. was supported in part by the National Institute of General Medical Sciences of the National Institutes of Health under Award Number T32GM008268. This work does not reflect the views of the funders. The funders were not involved in the design of the study, and funders do not have any ownership over the management and conduct of the study, the data, or the rights to publish. **Author contributions:** N.P. developed lyophilized RT-LAMP and outlined the overall plan for experiments and data visualization presented in this manuscript. R.G.A. developed a mobile software application for operating the device and analyzing results in real-time. M.R. developed the heater and fluorescent reader device. E.K. designed the RT-LAMP assay primers and probes and engineered polymerase. E.K., I.T.H., and Q.W. synthesized polymerase and provided usability feedback during the development of the Harmony COVID-19 system. I.T.H. and Q.W. prepared *in vitro* RNA transcripts. J.H.K. and V.L. led the usability study. N.P., A.K.O., and C.B. tested clinical and contrived specimens. R.G.A., N.P., A.K.O., and J.H.K. analyzed the data. I.T.H. and Q.W. prepared the kits and blinded mock samples for usability testing. D.L. developed housing units to integrate the cell phone, sample preparation station, and the device. L.M.S. and P.D.H. designed and characterized the clinical specimen panel used in this study. M.T. oversaw the usability study. B.R.L. oversaw the development and evaluation of the Harmony COVID-19 system. All authors contributed to writing this manuscript and approved the final version. **Competing interest:** Provisional patent applications have been filed on several components of the Harmony COVID-19 system. N.P., R.G.A., M.R., E.K., Q.W., I.T.H., D.L., and B.R.L. are inventors on one or more provisional patent applications. N.P., R.G.A., M.R., E.K., Q.W., I.T.H., J.H.K., A.K.O., C.B., D.L., and B.R.L. anticipate forthcoming equity in a startup company that has licensed related technology and supports ongoing work in the B.R.L. laboratory at the University of Washington but played no role in the study. M.T. is on the Advisory Board for Visby Medical which produces tests for COVID-19 and has received reimbursement for this work. He has also received reimbursement for medical advice to Roche Molecular Diagnostics and Inflammatix. **Data and materials availability:** All data are presented in the main text and supplementary information. **Code availability:** Sequence alignment and primer design was performed using Geneious 8.1.9 (Auckland, New Zealand). CFX-Maestro (BioRad, Hercules, CA) was used to analyze the RT-PCR results. TapeStation Software (Agilent, Santa Clara, CA) was used to visualize fragments of synthetic RNA. A provisional patent application was filed regarding aspects of the cell phone application. The data analysis algorithm is described in the manuscript.

## SUPPLEMENTARY INFORMATION

Trehalose can stabilize several enzymes, but we found that 2.5% (*w/v*) trehalose resulted in a slower reaction at 63°C, the optimal reaction temperature of the excipient-free RT-LAMP. An increase of reaction temperature to 65°C can speed up the trehalose-containing RT-LAMP reactions but resulted in unreliable detection at 20 RNA copies/reaction (**fig. S1**).

**fig. S1.**
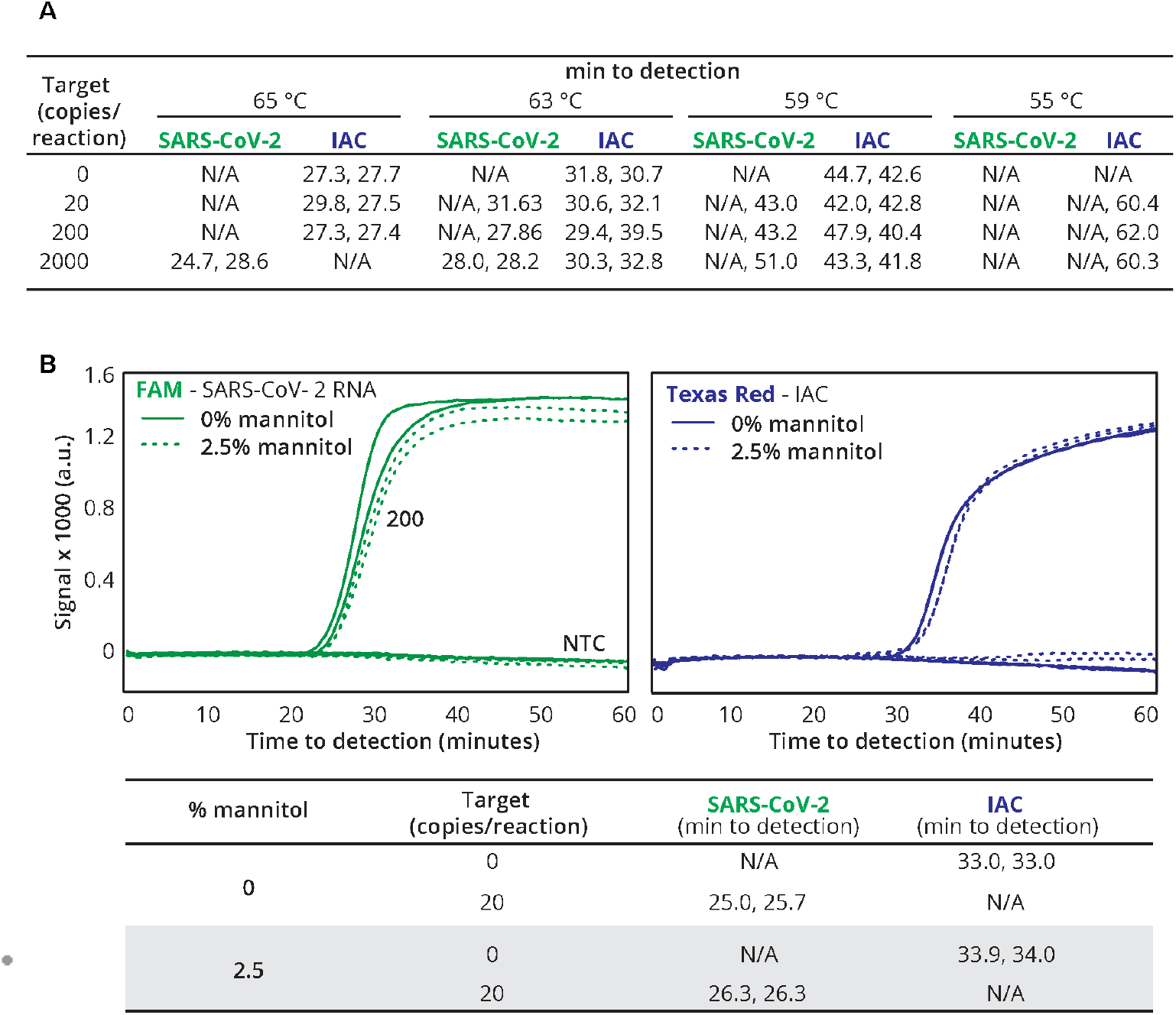
Effects of excipients on fresh RT-LAMP reactions tested on a commercial real-time thermal cycler. (**A**) Time to detection of SARS-CoV-2 RNA and IAC targets in the RT-LAMP containing 2.5% *(w/v)* trehalose and 0-2000 copies of SARS-CoV-2 RNA/40µL reaction. N/A means undetectable. For each condition, two technical replicates were run at 4 different reaction temperatures (i.e., 55°C, 59°C, 63°C, and 65°C). (**B**) Time to detection of SARS-CoV-2 RNA and IAC targets in the RT-LAMP reactions containing 0 or 20 copies of SARS-CoV-2 RNA/reaction, with and without 2.5% (*w/v*) mannitol, at 63°C.

**fig.S2.**
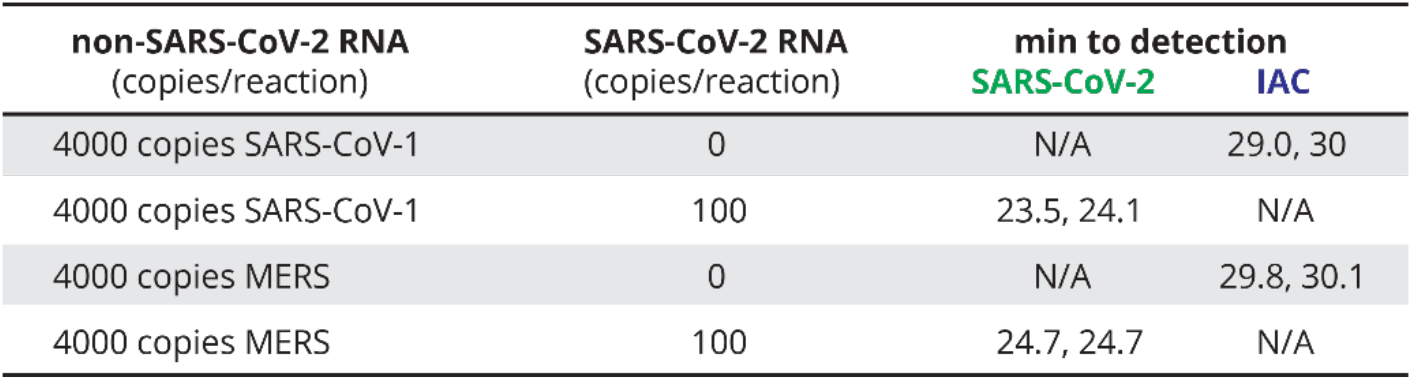
Cross-reactivity of MERS and SARS in lyophilized RT-LAMP. Detection time (min) of SARS-CoV-2 RNA and IAC targets in the presence of 4000 copies of either SARS-CoV-1 RNA or MERS RNA in 40µL RT-LAMP reactions at 63°C. Two technical replicates were conducted in each condition. In the reactions without SARS-CoV-2 RNA, we did not observe amplification of FAM signal but observed IAC amplification indicating the amplification reaction was functioning properly in each replicate. 100 copies of SARS-CoV-2 RNA were amplified in each positive control replicate, indicating proper function of SARS-CoV-2 amplification in this master mix.

**fig. S3.**
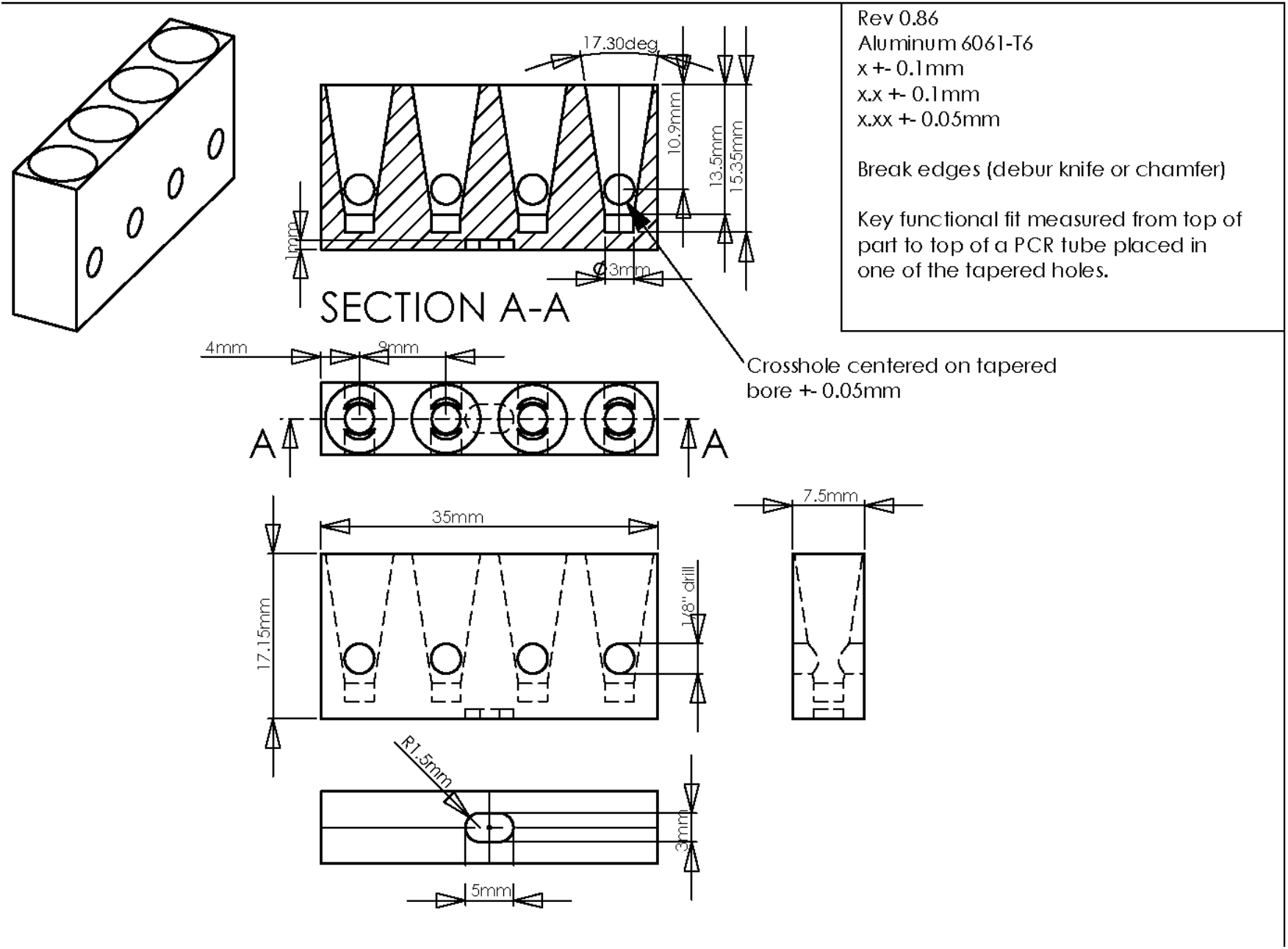
Heat block used in Harmony COVID-19 device. Drawing credit: Bryan Willman.

**fig. S4.**
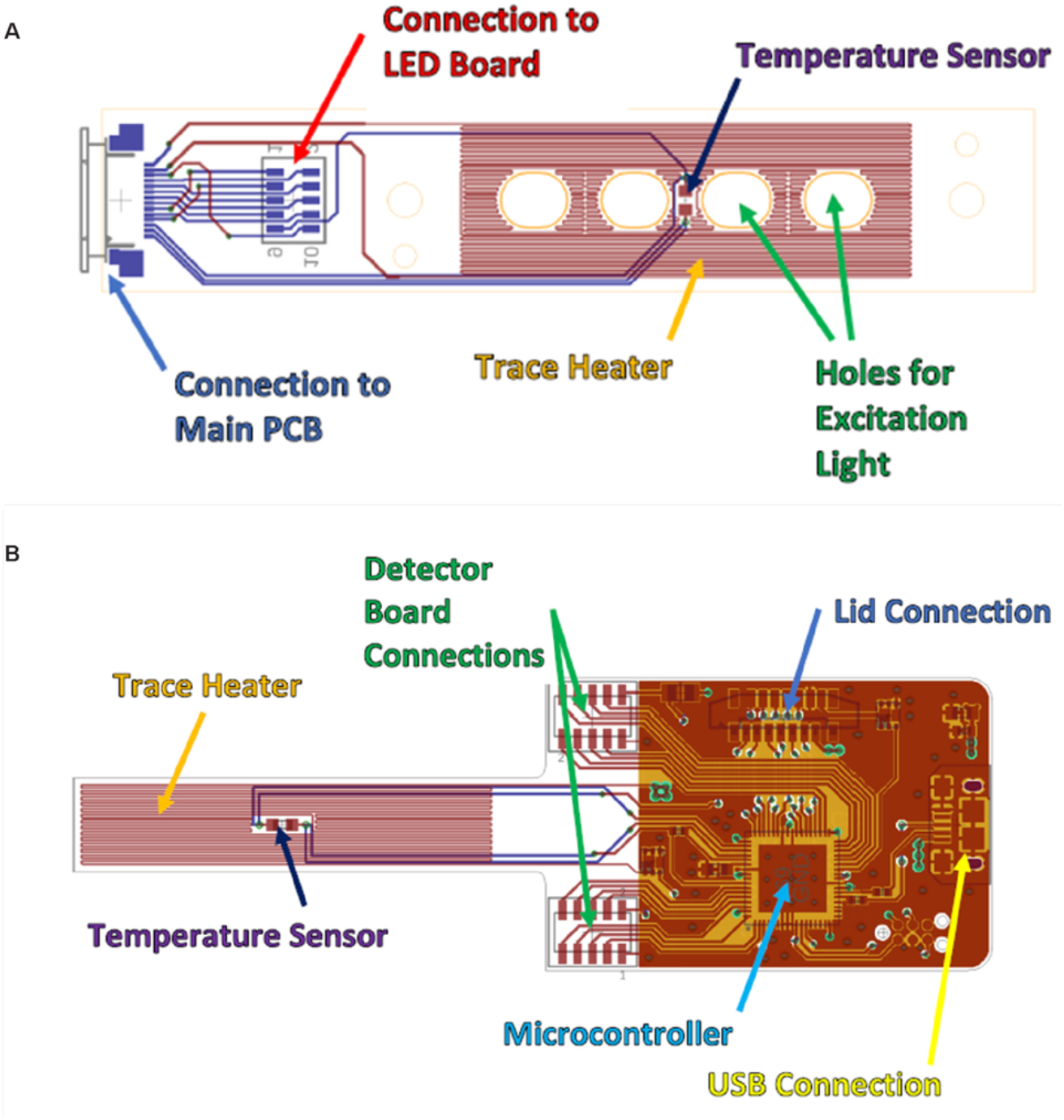
**The design of optical and heater circuit boards for the Harmony COVID-19 device**. (**A**) LED board (**B**) the detector board

**fig. S5.**
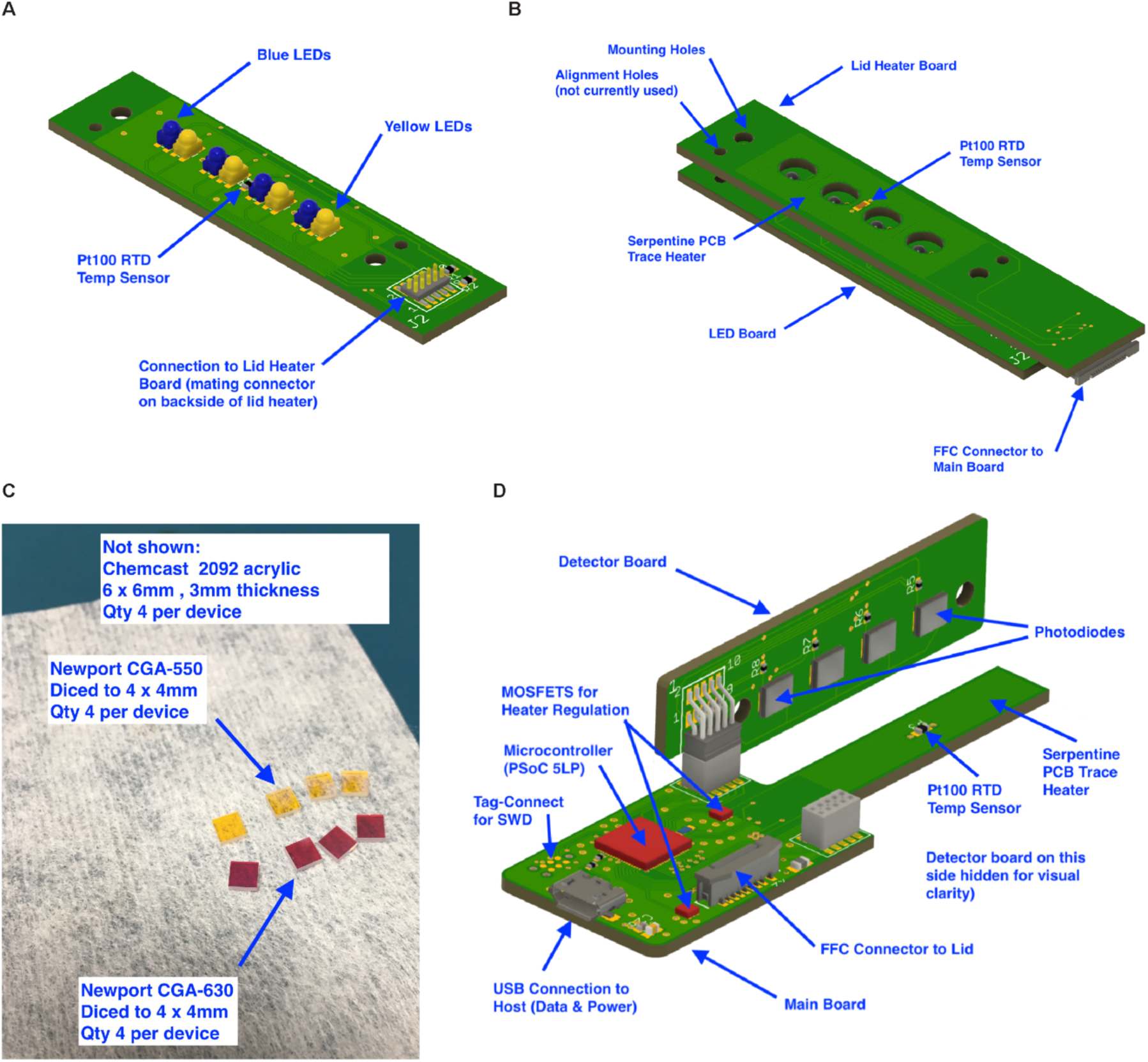
Circuit board assembly and required parts. (**A**) LED board. (**B**) LED board assembled to the lid heater board. (**C**) Custom-cut filter pieces. (**D**) Detector board and the main board. Each device require the following parts: thick film resistors (RCL12251R20FKEG, RCL12252R20FKEG, CRCW08050000Z0EBC and RCL12254R70FKEG from Vishey; RN73C1J75RBTDF from TE Connectivity; FTS-105-03-F-DV from Samtec; CR0402AJW-472GAS from Bourns), thin film resistors (RN73H1ETTP3003B25 from KOA Speer; ERJ-2RKF22R0X from Panasonic), multilayer ceramic capacitors (C0402C104M4RACAUTO, KEMET; 0603YC105KAT2A, AVX), multilayer ceramic capacitor (GRM188R60J226MEA0D, Murata), USB connectors (10118193-0001LF and 10118192-0001LF, FCI / Amphenol), FFC & FPC connector (52745-1497, Molex), standard yellow LEDs (LY E63B-CBEA-26-1-Z, Osram Opto Semiconductor), standard blue LEDs (150141BS63140, Wurth Elektronik), board-to-board & mezzanine connector (CLP-105-02-F-D and FLE-105-01-G-DV-K-TR, Samtec), FFC/FPC jumper cables (15166-0143, Molex), FFC & FFC connector (52559-1452, Molex), 2 headers & wire Housings (FTSH-105-01-F-DH, Samtec), MOSFET (FDMA410NZ, ON Semiconductor), 2 board-mount temperature sensors (PTS060301B100RP100 from Vishay), ARM Microcontrollers (CY8C5888LTI-LP097, Cypress Semiconductor), Photodiodes (SFH 2201, Opto Semiconductor).

**fig. S6.**
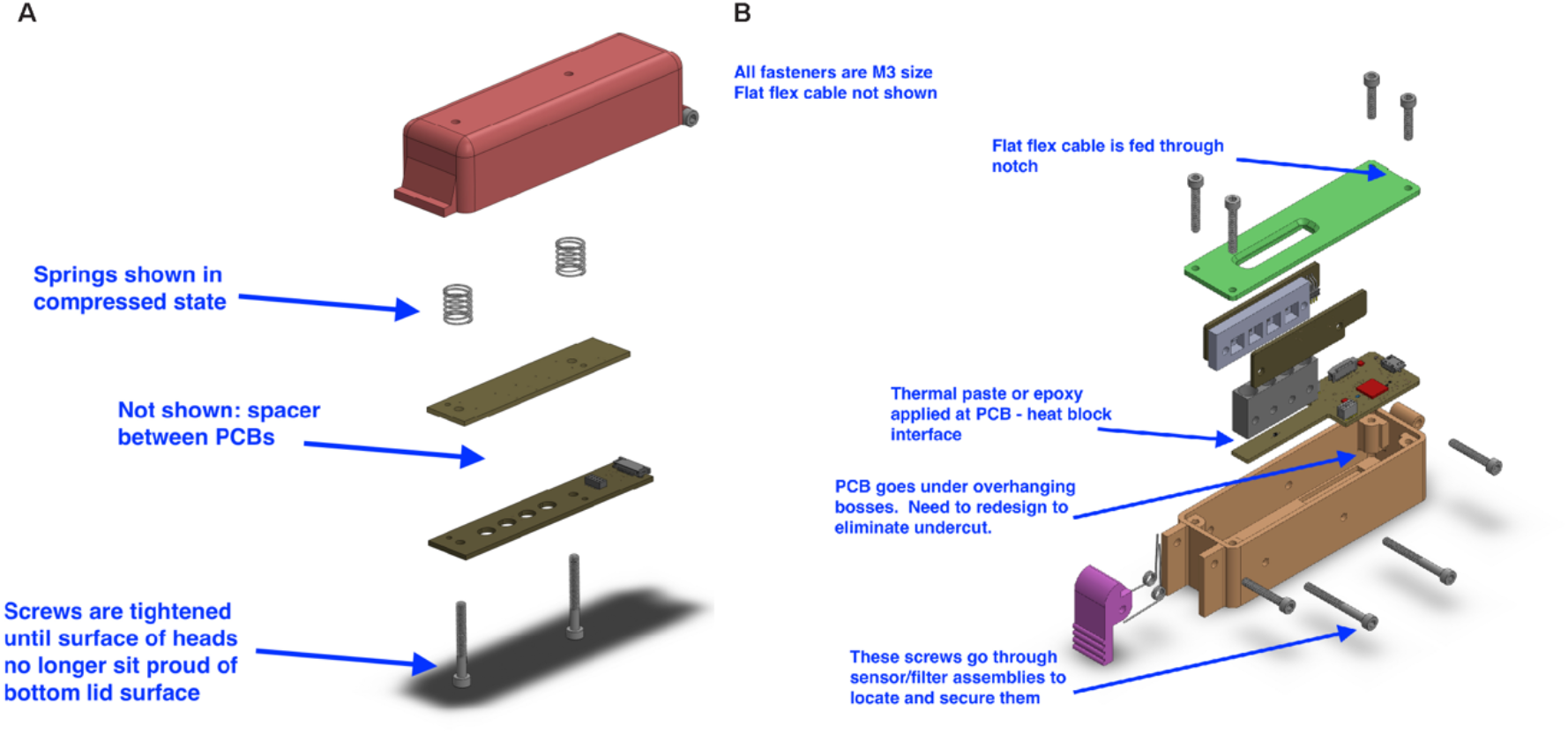
Device assembly. (**A**) Top part (**B**) Bottom part of the device are assembled using 10 screws, 2 compression springs, and 2 torsion springs (McMaster-Carr, Elmhurst, IL) including: Torsion Spring 90 Degree Angle, Left-Hand Wound, 0.204" OD (9271K109), Torsion Spring 90 Degree Angle, Right-Hand Wound, 0.204" OD (9271K143),Black-Oxide Alloy Steel Socket Head Screw M3 x 0.5 mm Thread, 5 mm Long (91290A110), Black-Oxide Alloy Steel Socket Head Screw M3 x 0.5 mm Thread, 10 mm Long (91290A115), Black-Oxide Alloy Steel Socket Head Screw M3 x 0.5 mm Thread, 15 mm Long (91290A572), Black-Oxide Alloy Steel Socket Head Screw M3 x 0.5 mm Thread, 20 mm Long (91290A123), Black-Oxide Alloy Steel Socket Head Screw M3 x 0.5 mm Thread, 25 mm Long (91290A125), Black-Oxide Alloy Steel Socket Head Screw M3 x 0.5 mm Thread, 30 mm Long, Partially Threaded (91290A130), Music-Wire Steel Compression Springs 0.875" Long, 0.36" OD, 0.296" ID (9434K73).

### video S1

Demonstration of transfer pipette workflow incorporated with Harmony COVID-19

### video S2

Demonstration of the integrated sampling system workflow with Harmony COVID-19 Both can be played from this slide presentation:

**table S1.**
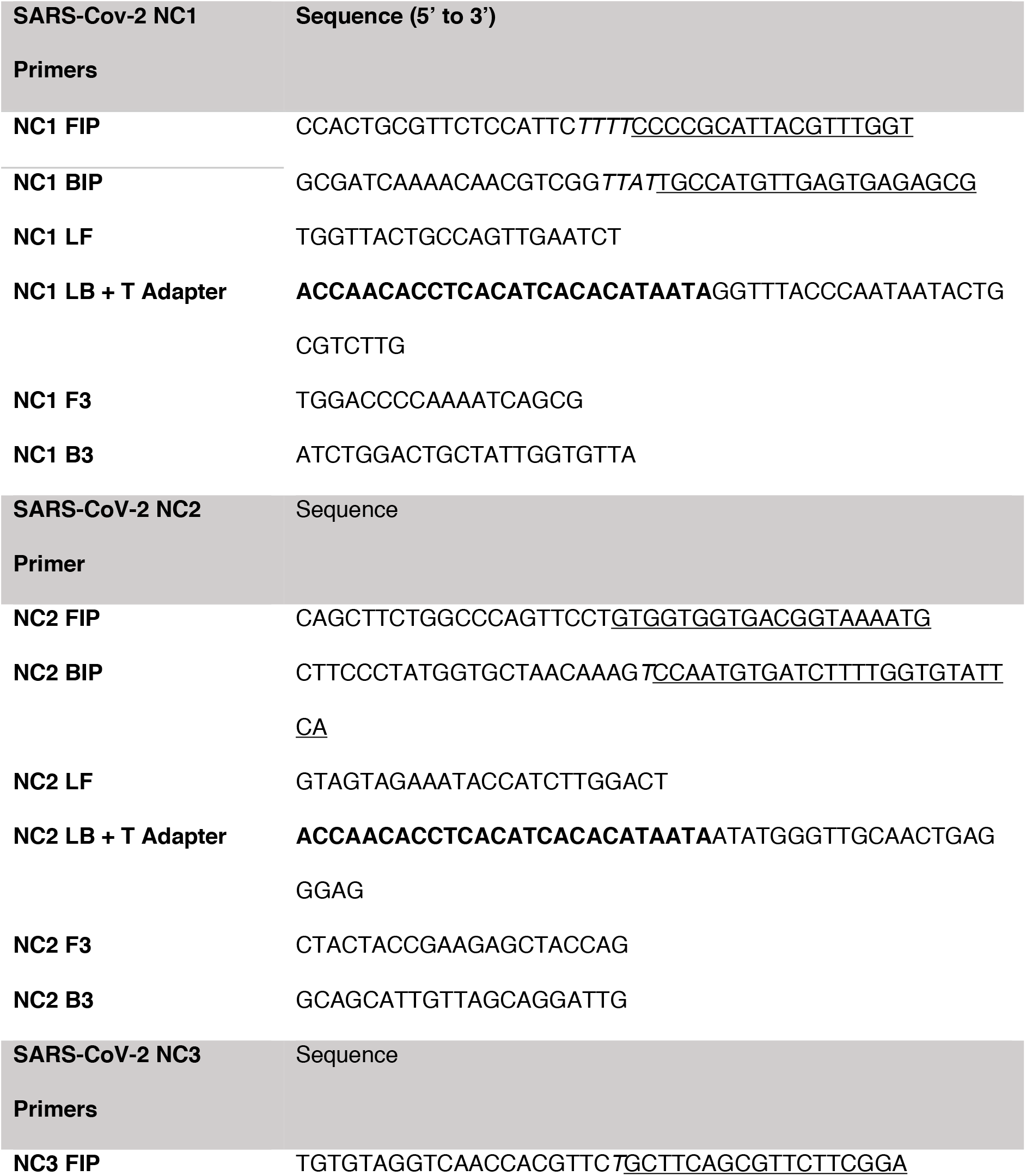
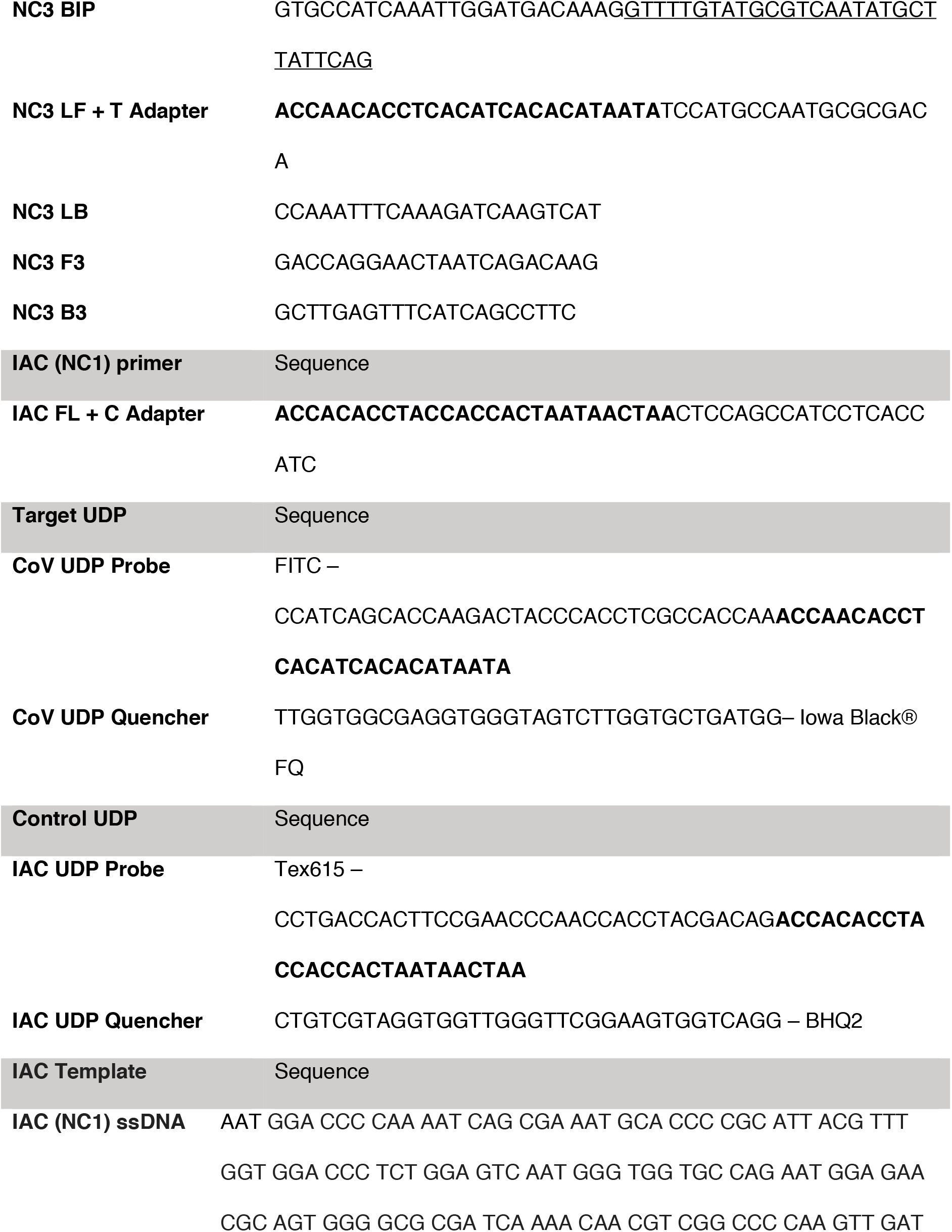
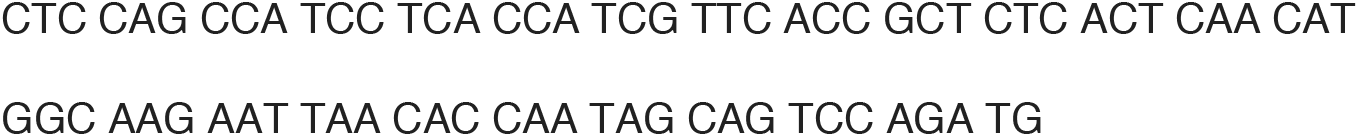
Primers, probes, and control sequences. For primers and probes F2/B2 sequences are underlined, non-template linker sequences are italicized, and adapter sequences are in bold.

### Sample transfer methods at the POC

Volumetric transfer pipettes are used in POC tests, but we have not identified a unified dispenser used in commercial pathogen tests. A unified dispenser integrating the swab and the buffer container in a single unit could reduce opportunities for sample mix-up when multiple samples are processed simultaneously. A simple workflow is crucial, especially in busy settings like clinics. However, in our hands, the in-house built dispenser led to variable dispensed fluid volumes and had a higher failure rate than those of the transfer pipette method. While the unified dispenser has many attractive features, we would not recommend using this in-house assembled dispenser unit until the method has been optimized to achieve a more accurately dispensed volume.

Among HCWs, 30% (3/10) reported problems dispensing the fluid using the unified system. However, 20% (2/10) of HCWs agreed that the unified dispenser offered an advantage in its similarity to other tools used in healthcare settings, and 30% (3/10) HCWs were concerned about contaminating the sample or the environment with the transfer pipette compared to the unified dispenser system. Only 1/10 (10%) HCW preferred the smallest tube (0.2mL) for either method, and 5/10 (50%) reported that larger (1.5mL) tubes were helpful for the unified system, while 40% (4/10) of HCWs said receptacle size did not make a difference when using the transfer pipette, and none reported preference for tube size when using the unified dispenser system. HCWs reported higher confidence in correctly completing the second kit compared to the first kit of each method, indicating a similar learning curve.

**table S2.**
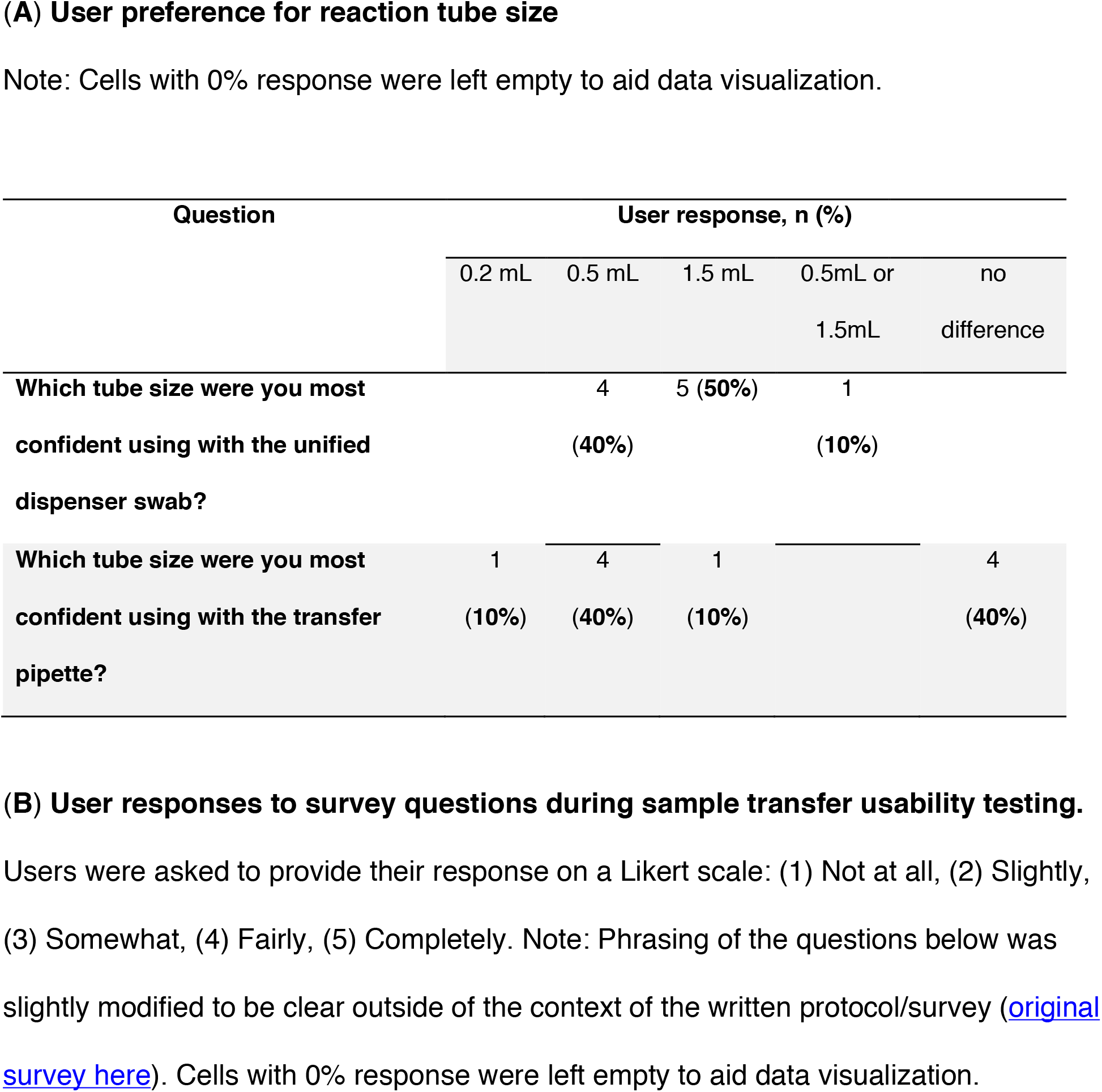
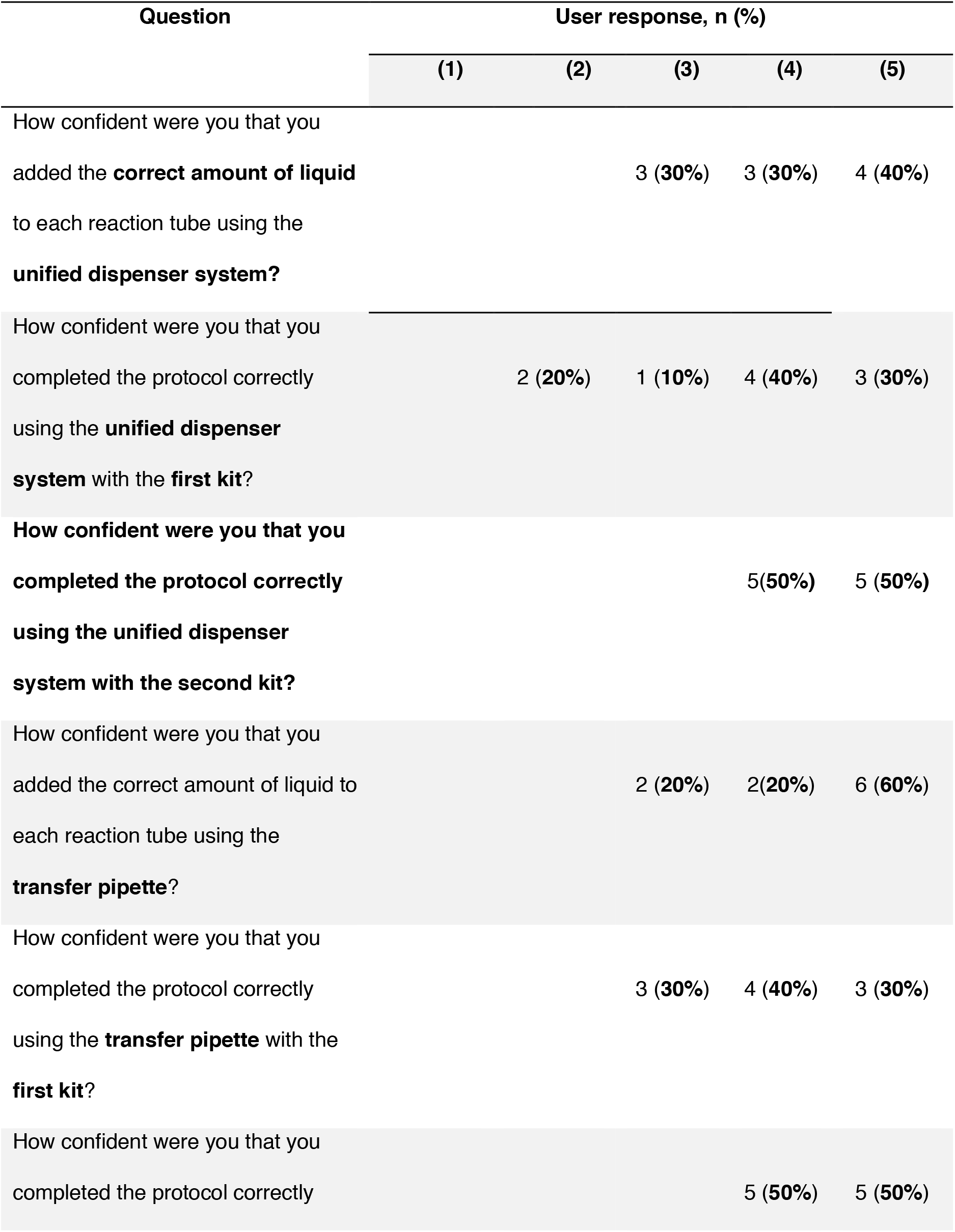
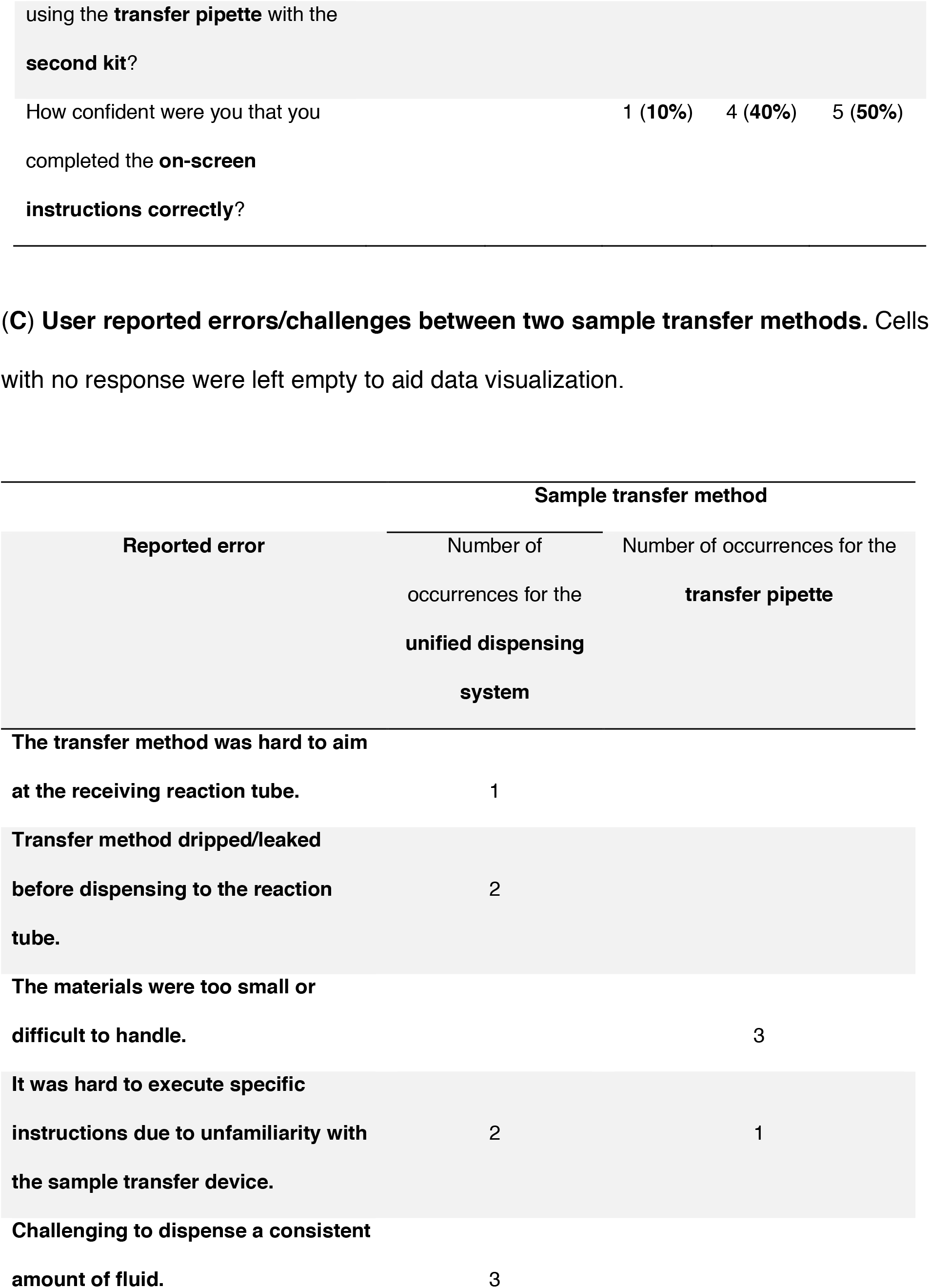
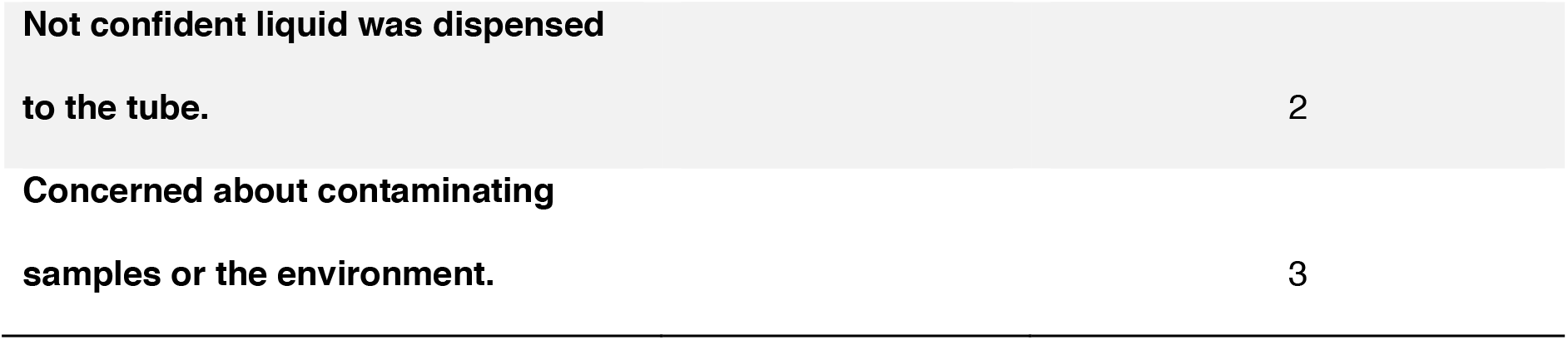
Feedback from the HCWs.

**table S3.** Detailed reagent and device cost at a production scale of 10,000 units.

| Item | Supplier | Cost/device (US\$) |
| --- | --- | --- |
| Detector boards | Macrofab | \$7 each x 2 (\$14 total) |
| Main board | Macrofab | \$25 each |
| LED board | Macrofab | \$9 each |
| Lid Heater | Oshpark | \$7 each |
| Heat block | Bryan Willman | \$12 each |
| Housing | Xometry – HP MultiJet<br>Fusion 3D printing, no<br>volume discount | \$50 (costs here should be reduced for<br>scale-up production) |
| Red filters | Newport | \$10 each, \$40 in total |
| Green filters | Newport | \$20 each, \$80 in total |
| Assembly | N/A | \$30 (half an hour assembly time at<br>\$60/hour cost) |
| Total | | \$267 per unit |

**table S4.** Consumable costs per test.

| Item | Supplier | Costs per kit (US\$) |
| --- | --- | --- |
| Sampling components<br>(\$2 maximum) | Multiple sources | \$2 |
| Desiccant | Multiple sources | \$0.5 |
| PCR tube | Multiple sources | Negligible |
| DNA polymerase | Produced in-house | Negligible |
| Reverse transcriptase | New England Biolabs | \$3.5 |
| Thermostable inorganic<br>pyrophosphatase | New England Biolabs | \$0.23 |
| Primers | Integrated DNA<br>Technologies | \$0.03 |
| Fluorescent<br>probe/quenchers | Integrated DNA<br>Technologies | \$0.24 |
| Triton-X100 | Sigma-Aldrich | Negligible |
| dNTPs | New England Biolabs | \$0.38 |
| Mannitol | OPS Diagnostics | Negligible |
| DL-Dithiothreitol (DTT) | Promega | Negligible |
| Nuclease-free water | VWR | Negligible |
| 1X Tris low-EDTA buffer | VWR | Negligible |
| Rnasin ribonuclease<br>inhibitor | Promega | \$0.71 |
| DNA internal<br>amplification control | Integrated DNA<br>Technologies | \$0.01 |
| **Cost of goods** | | **\$8.00** |

